# Spatially resolved transcriptomics of glomerular diseases

**DOI:** 10.64898/2026.09.23.26363804

**Authors:** Taehwan Shin, Stephan Weissbach, Paola Capodieci, Jeanine Henry-George, Christian Opitz, Samuel Barbieri, Xuedi Wang, Kristin Meliambro, Mark McConnell, Valerie Dubost, Eric Olson, Sophia Wild, Jia Jie Wu, Juliane Perner, Federico Tortelli, Joseph Loureiro, Paolo Cravedi

**Affiliations:** Disease Area eXploratory, Novartis Biomedical Research, Cambridge, MA, USA; Basel, Switzerland; Preclinical Safety, Novartis Biomedical Research, Basel, Switzerland; Discovery Sciences, Novartis Biomedical Research, Cambridge, MA, USA; Basel, Switzerland; Discovery/ Innovation/ Data Science AI Innovation Novartis Postdoctoral Fellowship Program and Postbaccalaureate Fellowship Program, Biomedical Education and Innovation, Novartis Biomedical Research, Novartis Pharma AG; Mt. Sinai, Translational Transplant Research Center, Department of Medicine, Icahn School of Medicine at Mount Sinai, New York, NY, USA

**Keywords:** Glomerulopathy, clinical spatial transcriptomics, histology-guided analysis, translational research, complement pathway

## Abstract

Understanding the shared and disease-specific mechanisms that drive glomerular injury remains a major unmet need for developing more precise and effective treatments for glomerular diseases. Here, we integrate histology-guided spatial transcriptomics with trajectory inference and cell-state deconvolution to resolve glomerular injury across Membranous Nephropathy, Focal Segmental Glomerulosclerosis, and Lupus Nephritis. Herein, we show that glomerular diseases arising from distinct initiating mechanisms converge on common pathways during progressive loss of kidney function. We used spatial transcriptomics to analyze kidney biopsies from 59 individuals with glomerular diseases and controls and identified 792 glomeruli, including structurally injured glomeruli missed by transcriptomic clustering alone. Glomeruli shared interferon- and complement-associated inflammatory programs together with suppression of oxidative and lipid metabolic pathways, while retaining disease-preferential transcriptional features. Increased pseudotime correlated with lower estimated glomerular filtration rate, and cell-state deconvolution linked progression to podocyte loss and cellular remodeling. Parallel analysis of passive Heymann nephritis, a rat model of membranous nephropathy, recapitulated complement activation and localized complement activity to multiple renal cell populations extending into the periglomerular niche. These findings define a spatially resolved, cross-disease continuum of glomerular injury and identify coordinated local complement activation as a convergent feature of progressive glomerular damage.

## INTRODUCTION

Glomerular diseases encompass disorders with distinct initiating mechanisms, histopathologic features, and clinical trajectories, and converge on progressive loss of filtration, glomerulosclerosis, and kidney failure. Membranous nephropathy (MN), focal segmental glomerulosclerosis (FSGS), and lupus nephritis (LN), for example, arise from different immune and cellular insults but can produce remarkably similar endpoints of chronic tissue injury (Lopez-Novoa, Rodriguez-Pena et al. 2011). This convergence suggests that, beyond disease-specific initiating events, glomerular disorders share downstream molecular programs that determine progression. Identifying these shared injury states, while distinguishing them from diagnosis-specific mechanisms, could provide a complementary framework for understanding why clinically distinct diseases progress toward common structural and functional outcomes.

Transcriptomic studies have substantially expanded our understanding of the cellular and molecular programs associated with kidney disease (Lake, Menon et al. 2023, Novella-Rausell, Grudniewska et al. 2023, Qing, Hu et al. 2025, Mao, Wei et al. 2026). However, bulk approaches average signals across anatomically and functionally distinct renal compartments, whereas single-cell and single-nucleus methods require tissue dissociation and thereby lose the spatial relationships that define kidney pathology (Fan et al., 2024; Hu et al., 2024; Huang et al., 2025; Janosevic et al., 2025). This limitation is particularly relevant to glomerular disease, in which injury is frequently focal or segmental and marked heterogeneity can exist among glomeruli within the same biopsy. Relatively preserved and severely injured glomeruli may coexist within a single tissue section, while molecular changes within the glomerular tuft may be coupled to immune, stromal, vascular, and tubular responses in the surrounding tissue. Consequently, analysis at the whole-biopsy level can obscure molecular states that occur only in subsets of glomeruli.

Here, we integrated histology-guided spatial transcriptomics with trajectory inference and reference-based cell-state deconvolution to characterize individual glomeruli from healthy kidneys and kidneys affected by MN, FSGS, or LN. Histopathologist-validated image analysis enabled the *in-silico* dissection of 792 glomeruli, including injured glomeruli incompletely recovered by transcriptomic classification alone. We asked whether glomeruli from etiologically distinct diseases remained molecularly segregated or instead occupied shared states along a common injury landscape. Across diseases, individual glomeruli distributed along overlapping but disease-biased trajectories characterized by progressive loss of podocyte and metabolic programs and increasing inflammatory and remodeling activity. Among the pathways identified independently across cross-sectional, histologic, trajectory, and intercellular-signaling analyses, complement pathway emerged as a prominent convergent feature of glomerular injury. We extended this analysis to passive Heymann nephritis, an experimental model of membranous nephropathy, using higher-resolution Visium HD profiling, and we identified complement-associated programs across glomerular and peri-glomerular cell populations in the preclinical model. Together, these analyses define a spatially resolved framework in which distinct glomerular diseases retain disease-preferential molecular features while converging on shared tissue states associated with loss of kidney function.

## RESULTS

### Patients’ characteristics

We analyzed kidney biopsies from 59 individuals, including 15 healthy controls, 13 patients with membranous nephropathy (MN), 13 with focal segmental glomerulosclerosis (FSGS), and 18 with lupus nephritis (LN) (**Fig. 1** and **Sup. Table 1**). LN patients were younger and disproportionally female as compared to MN, FSGS and CTRL groups **(Fig. 1a-c)**. The disease groups encompassed a broad spectrum of kidney dysfunction and proteinuria at the time of biopsy (before any immunosuppressive therapy initiation). Mean baseline eGFR was 55.9 ± 34.7 mL/min/1.73 m² in MN, 33.7 ± 17.3 mL/min/1.73 m² in FSGS, and 53.0 ± 27.1 mL/min/1.73 m² in LN, whereas median proteinuria was 4.2, 2.2, and 1.1 g/day, respectively, and baseline creatinine levels are similar across groups **(Fig. 1d-f)**. Longitudinal follow-up further demonstrated substantial heterogeneity in kidney function trajectories. MN had the most favorable mean eGFR slope (0.7 ± 2.9 mL/min/1.73 m²/year), whereas FSGS showed the greatest average decline (-3.9 ± 4.7 mL/min/1.73 m²/year); LN showed greater interindividual variability, with a mean slope of -0.4 ± 11.1 mL/min/1.73 m²/year (**Fig. 1g,h**).

**Figure 1.**
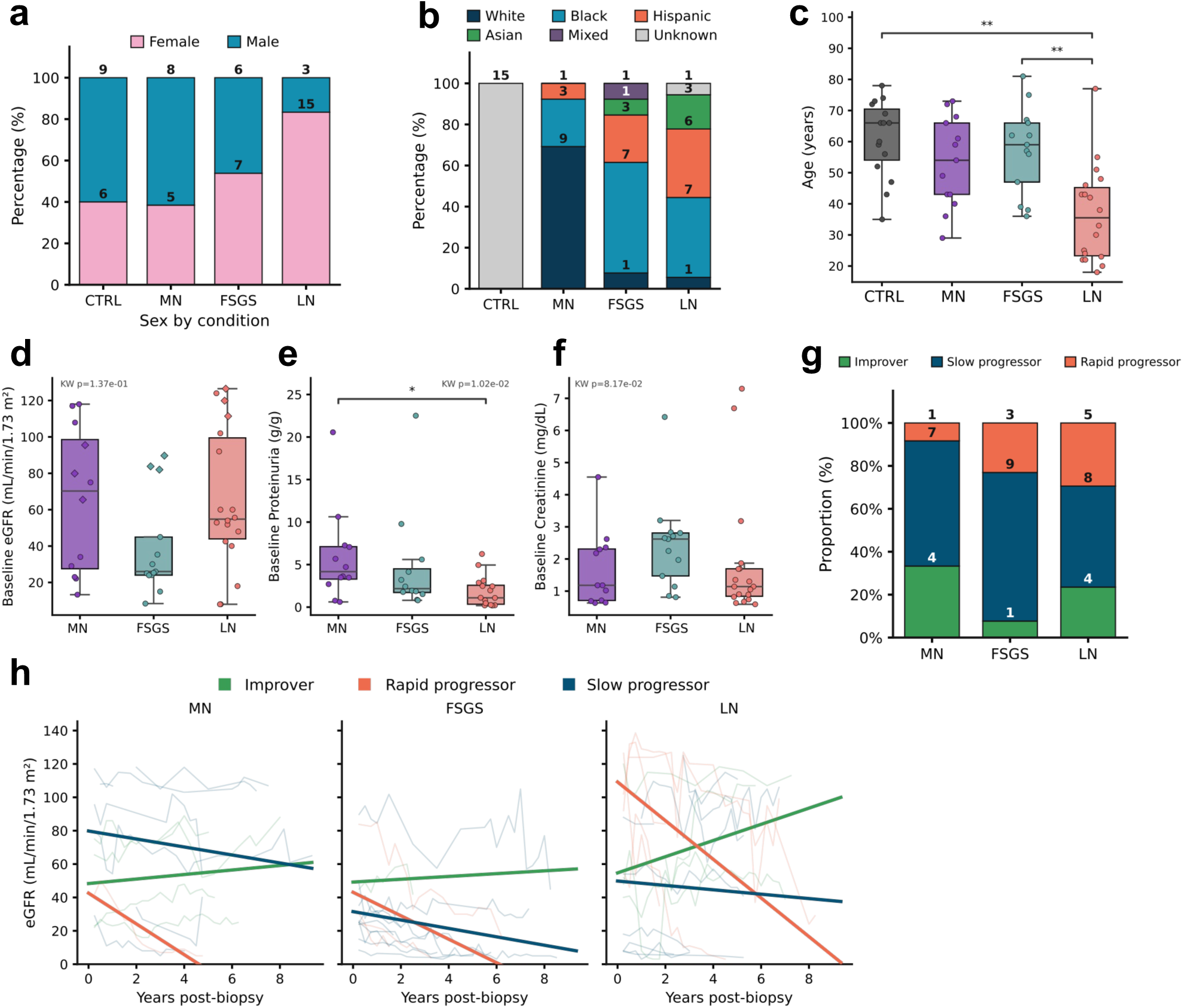
Clinical and demographic characteristics of the patient cohort by disease group. (**a**) Sex distribution, (**b**) self-reported ancestry, and (**c**) age at biopsy. (**d**) Baseline eGFR (mL/min/1.73 m²), (**e**) Baseline proteinuria (g/day), and (**f**) serum creatinine (mg/dL) at biopsy. (**g**) Proportion of eGFR progressor classes (Improver, Slow progressor, Rapid progressor) derived from linear mixed-effects model slope estimates. (**h**) Individual patient eGFR trajectories over time post-biopsy, faceted by disease group and colored by progressor class. Group differences in (c–f) were assessed by Kruskal-Wallis test across all groups; brackets indicate pairwise Mann-Whitney U comparisons (*p < 0.05, **p < 0.01).

### Histology-guided spatial transcriptomics captures heterogeneous glomerular injury across distinct diseases

We next generated spatial transcriptomic profiles from the biopsy specimens and initially identified glomerular regions using unsupervised transcriptomic clustering and canonical marker expression (**Fig. 2a-e**). Visual comparison with the corresponding histology, however, revealed two complementary sources of error: inclusion of spots extending beyond the glomerular boundary and failure to identify histologically recognizable glomeruli with attenuated glomerular transcriptional signatures **(Fig. 2b,f,g)**. We therefore trained a HALO AI classifier to identify glomeruli directly from the H&E images and subjected the resulting annotations to histopathologist review. This histology-guided approach both refined glomerular boundaries and recovered glomeruli missed by transcriptomic classification alone, enabling the *in-silico* dissection of 792 individual glomeruli across the cohort. The discrepancy was most evident in structurally injured and sclerotic glomeruli, particularly in FSGS, indicating that reliance on transcriptional identity alone preferentially excludes the glomeruli with the greatest remodeling.

**Figure 2.**
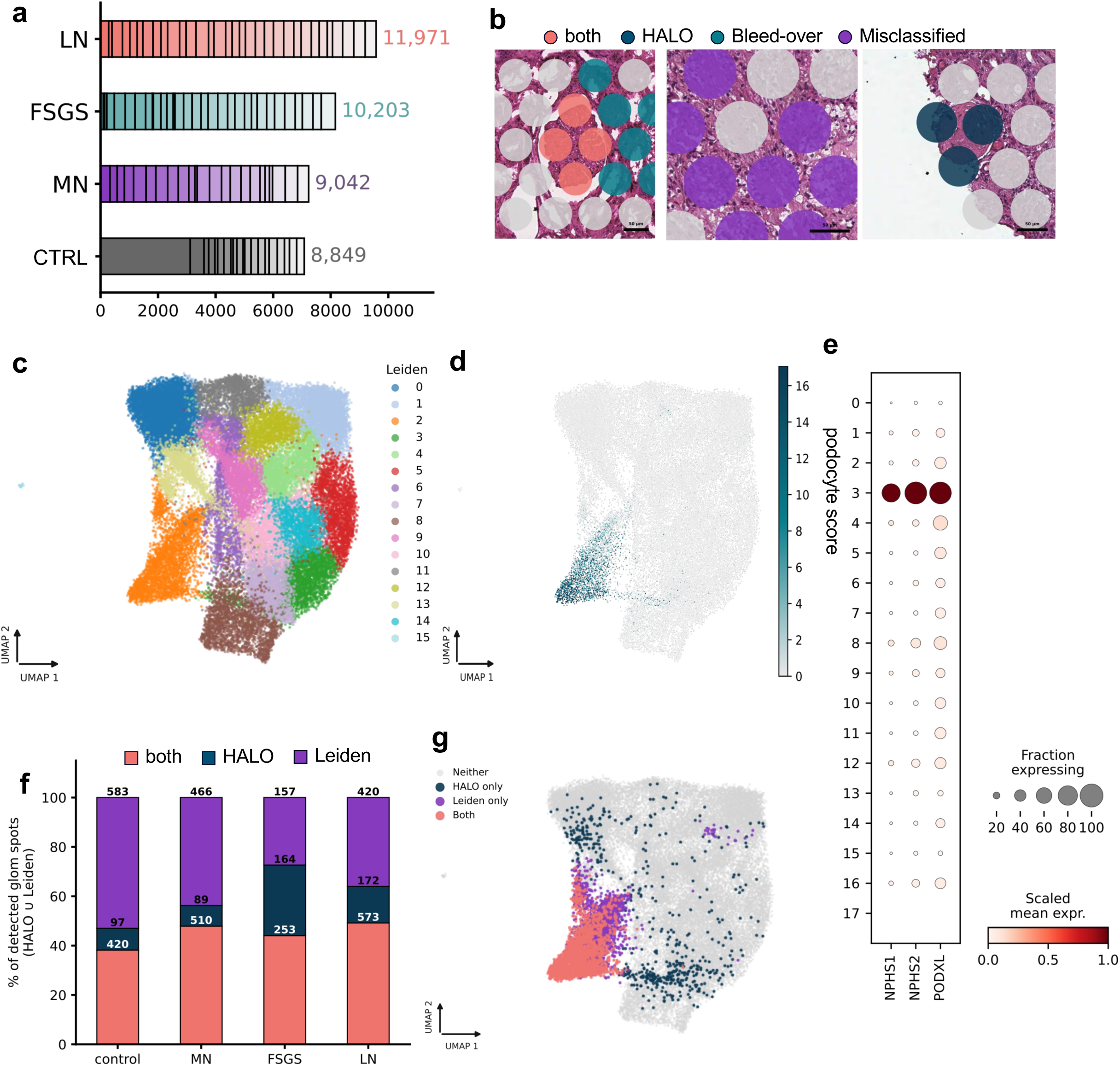
Visual and transcriptional demarcation of glomeruli. (**a**) number of 55 um spots with detectable gene expression available for the identification of glomeruli, (**b**) visual inspection of HALO AI predictions identified both type I and type II errors resolved by visual adjudication. (**c,d,e**) Leiden clustering of spots identified one with distinct podocyte score (Leiden 2, orange) shown in podocyte score heatmap (**d**) and using marker genes (**e**). (**f,g**) visual and transcriptional detection of glomeruli are complementary approaches that increase accuracy and precision of transcriptional analysis.

To independently assess the biological severity of the captured glomerular states, individual glomeruli were ranked histologically for injury and sclerosis by a histopathologist blinded to the sample origin (**Fig. 3**). Controls and MN contained the largest proportions of morphologically preserved glomeruli, whereas FSGS and LN were enriched for injured glomeruli, with FSGS showing a greater representation of advanced injury and sclerosis (**Fig. 3a,b,d,e**). Importantly, increasing histologic injury was associated with progressively poorer recovery of glomeruli by transcriptomic clustering, providing direct evidence that histology-guided annotation prevents systematic loss of advanced disease states. At the patient level, mean glomerular injury score correlated inversely with baseline eGFR (R = -0.45, P < 0.01), but not with subsequent eGFR slope (R = -0.07, P = n.s.)**(Fig. 3a,c; Sup. Fig. CLINIC d)**.

**Figure 3.**
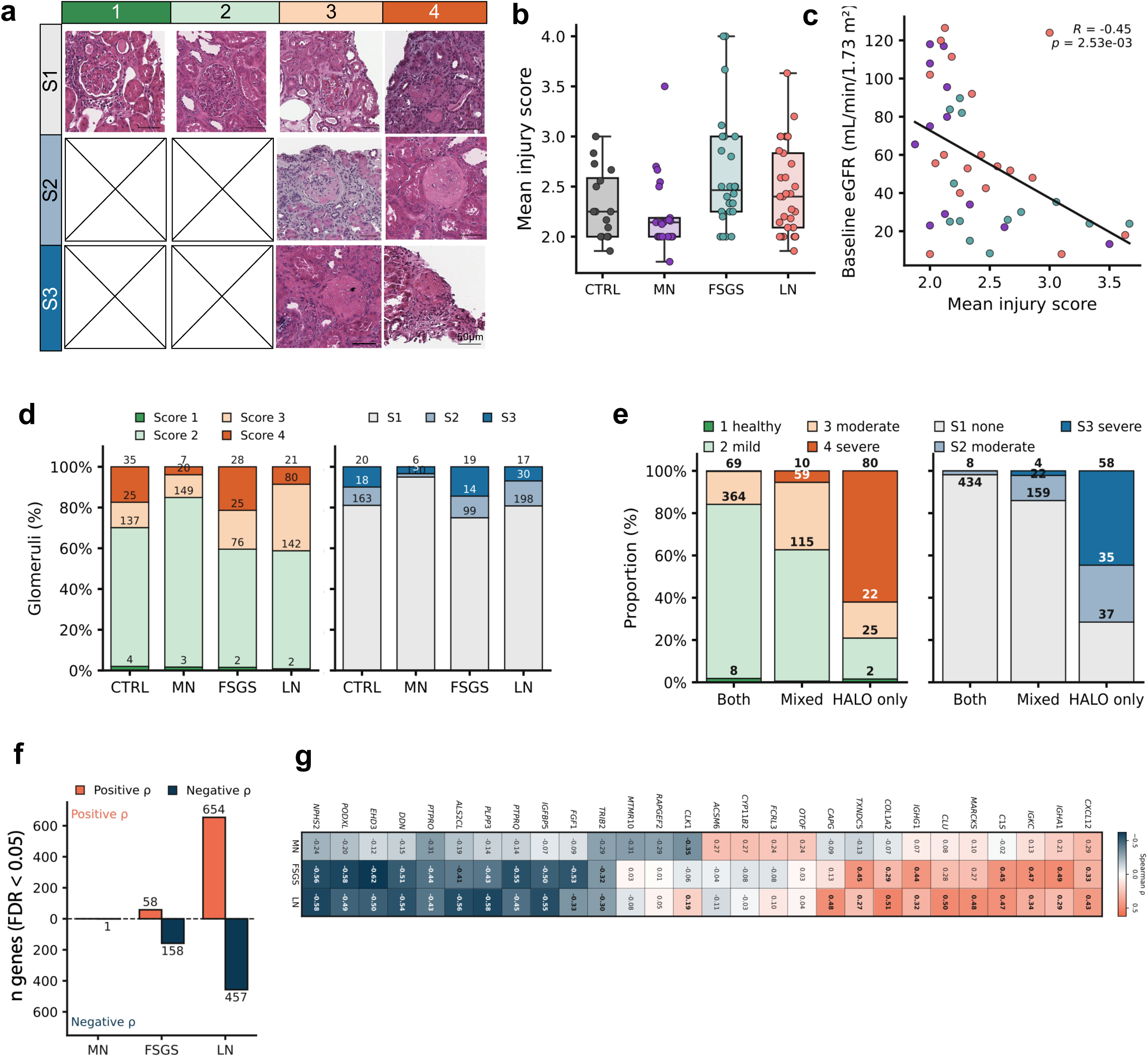
Glomerular injury and sclerosis scoring. (**a**) Representative histological images of glomeruli grouped by injury score (1-4, columns) and sclerosis score (S1–S3, rows). (**b**) Distribution of injury score, **(c)** baseline eGFR versus mean per-patient injury score, colored by disease condition, with linear regression fit. Regression line show Pearson correlation coefficient (R) and p-value. (**d**) Distribution of injury and sclerosis scores by disease group, (**e**) Injury score (left) and sclerosis score (right) by glomerulus detection method - fully captured by transcriptomic clustering and HALO (Both), partially captured (Mixed), or HALO-only. (**f**) Number of genes significantly correlated with ordinal injury group (Low/Moderate/Severe) per disease condition, split by direction of correlation. (**g**) Heatmap of Spearman ρ between top correlated genes and ordinal injury group, significant genes correlations are bold. Gene-injury group correlations were computed by Spearman correlation with Benjamini-Hochberg FDR correction; significance threshold FDR < 0.05.

Histologic injury was also accompanied by marked transcriptional remodeling. Only one transcript was significantly associated with injury in MN, compared with 216 in FSGS and 1,111 in LN **(Fig. 3f)**. Increasing injury was consistently associated with loss of podocyte and filtration-barrier transcripts, including *NPHS2*, *PODXL*, *PTPRO*, and *DDN*, together with reduced *EHD3* (glomerular endothelia cells), *C1S*, and the matrix-associated gene *COL1A2* increased with injury, particularly in FSGS and LN **(Fig. 3g)**. Thus, histologic deterioration was coupled to loss of glomerular cellular key features and acquisition of inflammatory and matrix-remodeling programs.

### Distinct glomerular diseases converge on shared inflammatory and metabolic programs

We next asked whether glomerular transcriptional remodeling reflected diagnosis-specific mechanisms or common responses to injury. Differential expression analysis was therefore performed using patient-level pseudobulk profiles generated exclusively from histology-defined glomeruli (**Fig. 4; Sup. Table 2**). All three diseases differed from healthy controls, although the magnitude and heterogeneity of the response varied substantially **(Fig. 4a-c)**. FSGS and LN showed the most extensive transcriptional remodeling, whereas MN displayed more modest changes and greater overlap with controls. Principal component analysis showed broad dispersion of FSGS glomerular profiles and separation of LN from controls, while MN occupied a more heterogeneous region overlapping both healthy and diseased states **(Fig. 4g)**.

**Figure 4.**
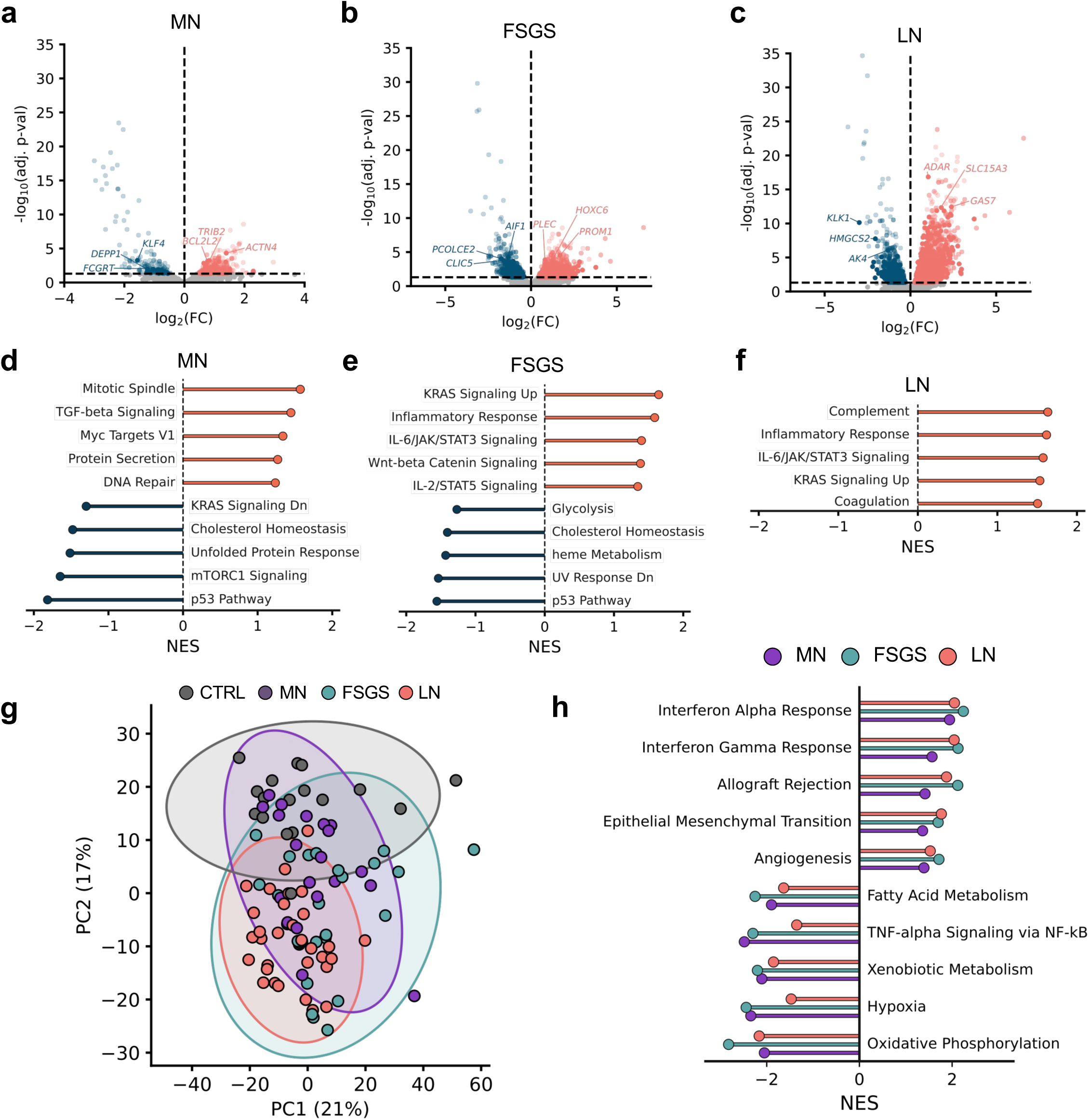
Differential gene expression and pathway analysis across disease groups. (**a–c**) Volcano plots of differential gene expression in pseudobulked glomerular transcriptomes per patient for (**a**) MN, (**b**) FSGS, and (**c**) LN. Points are colored by direction of change (up/downregulated); semi-transparent points denote genes also differentially expressed in at least one other disease group, opaque points denote genes uniquely differentially expressed in that disease group. (**d-f**) Gene Set Enrichment Analysis (GSEA, MSigDB human 2025) of top up- and downregulated pathways for (**d**) MN, (**e**) FSGS, and (**f**) LN, restricted to pathways not concordantly enriched (same direction) across all three disease groups. (**g**) Principal component analysis of pseudobulked glomerular transcriptomes per patient, colored by condition (CTRL, MN, FSGS, LN). Ellipses represent 95% confidence regions for each group, (**h**) GSEA pathways significantly enriched in all three disease groups (MN, FSGS, LN), shown as normalized enrichment score (NES) per group. Differential expression was assessed by DESeq2 (Wald test) on pseudobulked glomerular expression per patient; dashed lines in (**b-d**) indicate the adjusted p-value significance threshold. GSEA results in (**e-h**) are shown as NES, filtered at FDR < 0.25, top 5 pathways.

Pathway analysis reinforced this convergence. Interferon-α response, interferon-γ response, and allograft-rejection gene sets were concordantly enriched across MN, FSGS, and LN, whereas oxidative phosphorylation and fatty-acid metabolism were consistently reduced **(Fig. 4h)**. Thus, despite distinct etiologies, all three diseases shared a transcriptional state characterized by inflammatory activation accompanied by suppression of metabolic homeostasis. Superimposed on this common response were disease-preferential programs. MN showed enrichment of TGF-β signaling, mitotic-spindle, and MYC-target programs together with reduced KRAS signaling and cholesterol homeostasis **(Fig. 4d)**. FSGS was characterized by inflammatory and IL-6/JAK/STAT3 programs together with altered KRAS signaling and .pression of glycolytic and cholesterol-homeostasis pathways **(Fig. 4e)**. LN showed particularly prominent inflammatory, IL-6/JAK/STAT3, and complement enrichment, and no downregulated gene sets **(Fig. 4f)**. These findings defined a molecular architecture in which diagnosis-specific programs coexist with a narrower set of shared inflammatory and metabolic responses.

### Individual glomeruli converge along shared but disease-biased injury trajectories

We next asked whether individual, spatially defined glomeruli could be ordered along common trajectories of injury, irrespective of clinical diagnosis **(Fig. 5)**. Application of Slingshot trajectory inference to histology-defined glomerular transcriptomes identified five lineages spanning the transcriptional landscape of these kidney biopsies **(Fig. 5a**). Pseudotime lineages projected on UMAP plot of spatially defined glomeruli have distinct representations of patient groups. Lineages 3 and 5 were preferentially populated by control glomeruli **(Sup. Fig. PSEUDOTIME d)**, whereas Lineages 1, 2, and 4 contained greater proportions of disease-associated glomeruli **(Fig. 5b)**. Thus, the trajectories did not simply reproduce diagnostic categories, but instead resolved overlapping disease states within a common transcriptional space **(Fig. 5b,c)**.

**Figure 5.**
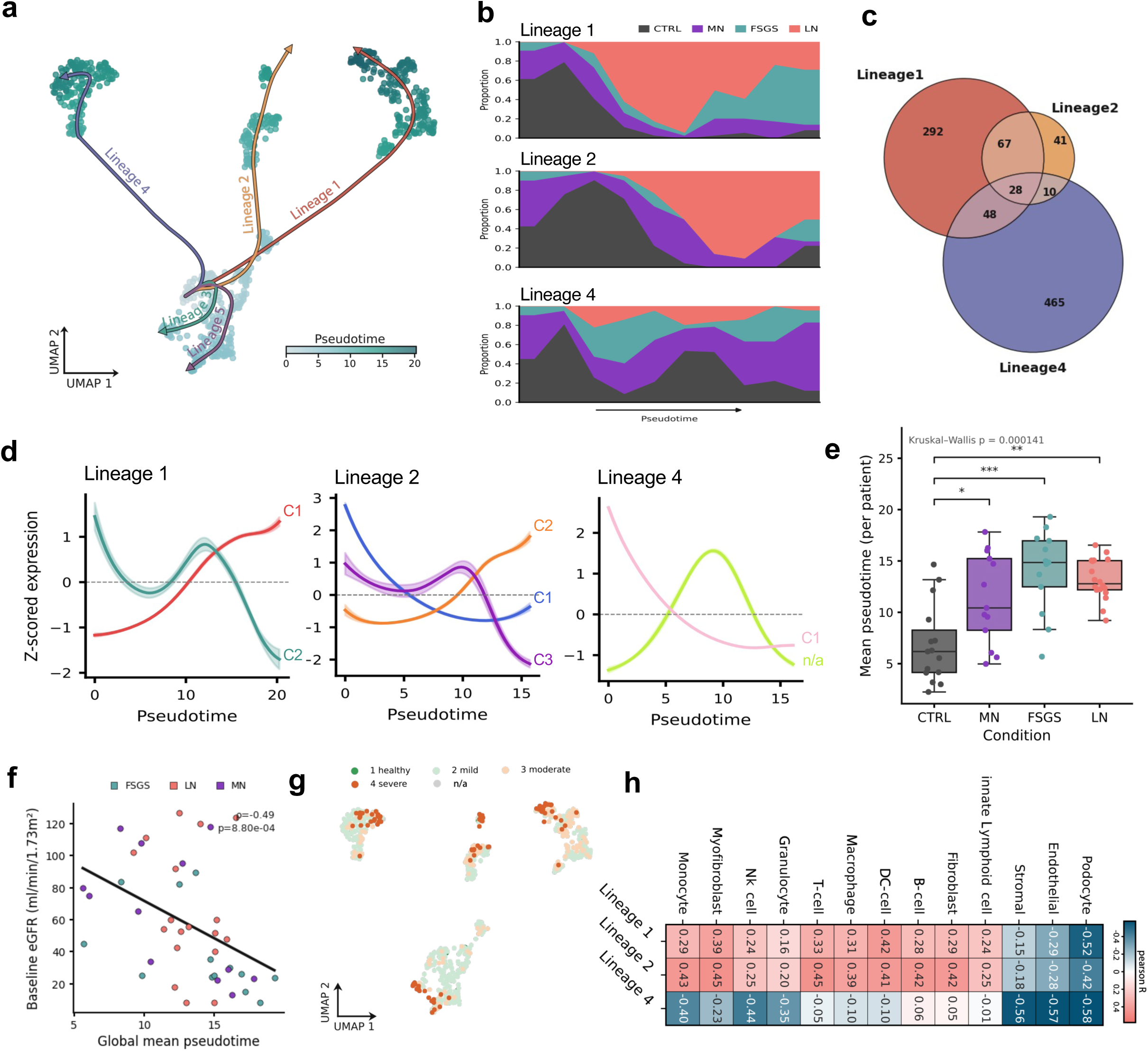
Pseudotime trajectory analysis of glomerular transcriptomes. (**a**) UMAP of pseudobulked glomerular transcriptomes with Slingshot trajectory analysis, showing 5 inferred lineages, colored by pseudotime. (**b**) Disease composition along pseudotime for Lineages 1, 2, and 4. (**c**) Overlap of significantly enriched pathways (ORA, FDR < 0.05) across Lineages 1, 2, and 4. (**d**) Hierarchical clustering of genes significantly associated with pseudotime, shown as smoothed Z-scored expression per cluster, for Lineages 1, 2, and 4. (**e**) Mean pseudotime values patient glomeruli per condition. **(f)** pearson correlation between baseline eGFR and global mean pseudotime value. **(g)** UMAP colored by histopathologist-assigned glomerular injury score (1 healthy, 2 mild, 3 moderate, 4 severe; n/a = unscored). (**h**) Celltype enrichment across the three lineages. Enrichment assessed by over-representation analysis (ORA); asterisks in (e) denote FDR-adjusted significance (*p < 0.05, **p < 0.01, ***p < 0.001, ****p < 0.0001).

Independent histologic assessment supported the biological ordering of these trajectories **(Fig. 5d,e)**. Diseased kidneys were shifted toward later injury states compared with controls (**Fig. 5e)**, but glomeruli occupying relatively preserved and advanced states frequently coexisted within the same biopsy **(Sup. Fig. PSEUDOTIME2)**. Despite this intratissue heterogeneity, the mean glomerular pseudotime of each biopsy was inversely associated with baseline kidney function (ρ = -0.49, P < 0.001; **Fig. 5f**), associating the transcriptional position of individual glomeruli along the inferred injury continuum to measurable loss of kidney function.

Histopathologist-assigned injury scores projected onto the trajectory map order morphologically preserved glomeruli preferentially to earlier states and glomeruli with moderate or severe injury accumulated toward lineage endpoints **(Fig. 5g)**. The computational trajectories therefore captured a diversity of glomeruli states, and we next modeled pathway inferences along each lineage using tradeSeq. The magnitude of transcriptional remodeling varied markedly among trajectories, ranging from 36 pseudotime-associated genes in Lineage 3 to 2,823 in Lineage 4 **(Sup. Fig. PSEUDOTIME2 a)**. Despite this heterogeneity, pathway analysis identified considerable overlap among injury-associated gene clusters. Increasing expression along Lineages 1 and 2 was associated with complement, interferon response, allograft-rejection, and epithelial-to-mesenchymal-transition programs, revealing a recurrent inflammatory and remodeling response across trajectories with different disease compositions **(Sup. Fig. PSEUDOTIME2 b)**. Conversely, the dominant Lineage 4 program was characterized by progressive loss of podocyte structural genes, including *NPHS1*, *NPHS2*, *SYNPO*, and *ITGB1*, together with suppression of oxidative phosphorylation, electron-transport, cytoskeletal, and cell-survival programs.

### Cellular remodeling and coordinated inflammatory signaling identify complement as a convergent injury program

We next asked whether glomerular injury progression was accompanied by changes in cellular composition. Because individual Visium spots contain transcripts from multiple neighboring cells, we deconvolved spatial profiles using the Kidney Precision Medicine Project (KPMP) single-nucleus reference atlas (Lake, Menon et al. 2023). Predicted podocyte abundance closely colocalized with histologically defined glomeruli and correlated with visual glomerular annotation across disease groups, with correlations of approximately 0.62-0.65 in CTRL, MN, and LN and a modestly lower correlation of 0.56 in FSGS (**Sup. Fig. CELLDECON a**). Cell-type deconvolution revealed disease-dependent remodeling within glomerular regions. Podocyte abundance was reduced in FSGS and LN compared with controls (**Sup. Fig. CELLDECON b**). Correlations further indicate systematic compositional changes across the injury continuum, with podocyte and myofibroblast states strongly pseudotime-associated to compartments **(Sup. Fig. CELLDECON c,d,e)**. These findings indicate that injury progression reflects a remodeling of the cellular composition of the glomerulus.

Multiple analytical approaches indicate transcriptional modulation of the complement pathway across the three diseases, which we analyze in detail along lineage 1 where all four sample types are present **(Fig. 6)**. We divided the glomeruli in pseudotime lineage 1 into tertiles **(Fig. 6a)**, and we measured complement pathway activity within and adjacent to each glomerulus **(Fig. 6b)**. When we examined expression differences of these pathways along spatial distance to the glomeruli, we observed the highest expression of the pathway in glomerulus and decreasing markedly with distance from the glomerular boundary. LN had pronounced complement pathway followed by FSGS, while MN showed more significant distal/non-glomerular enrichment (**Fig 6b**).

**Figure 6.**
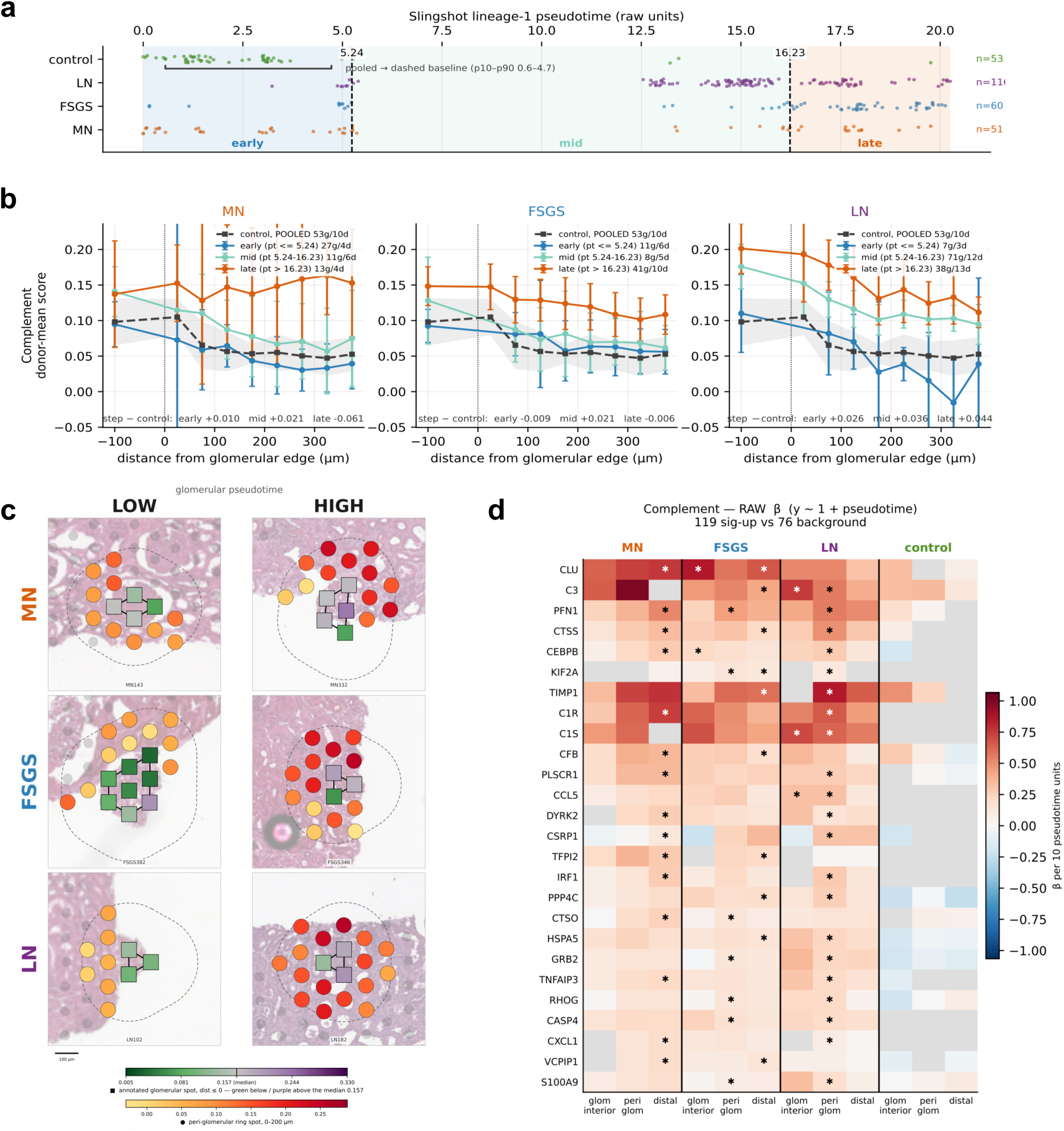
Evidence for endogenous complement pathway activation in human glomeruli. (**a**) distribution of glomeruli across lineage 1 faceted by disease group, split into tertiles at dashed lines, (**b**) plots of complement pathway modulation in the three disease groups separated by lineage 1 tertiles show distance for glomerulus center with boundary set at zero. The dashed black curve with the grey 95% ribbon is the pooled control baseline. *x* is the signed distance to the glomerulus’ hull edge, binned (−200–0 = "interior", then 50 µm bins to 400 µm) and plotted at the bin centre; the axis stops at 400 µm by construction. *y* is the **donor mean**: the score is averaged over the spots of one donor inside a (band × distance bin) cell first, and the plotted point is the mean over donors with a *t*-based **95 % CI across donors** — so the effective *n* of every point is its donor count, printed per band in the legend as Ng/Nd (MN 27/4, 11/6, 13/4; FSGS 11/6, 8/5, 41/10; LN 7/3, 71/12, 38/13). The number under each panel is the descriptive **interior→375 µm step minus the control step** in the same units. (**c**) Visualization of complement pathway expression in individual glomeruli. Rows are conditions, columns are the low and high shared bands. Each panel is **one** glomerulus — the one nearest that cell’s median pseudotime among those with ≥ 5 ring spots — drawn on its own section’s H&E, all six at one magnification (±368 µm). Squares are annotated glomerular spots on a diverging green–purple scale split at the median annotated-spot score; circles are the 0–200 µm peri-glomerular ring on a sequential yellow–red scale. The solid outline is the glomerulus’ convex hull of annotated spot centres (the distance = 0 edge) and the dashed outline is that hull buffered by 200 µm (the ring’s outer edge, verified spot-by-spot against the tabulated distances). (**d**) **Per-gene, per-compartment association with L1 pseudotime.** Rows are the top 26 of 83 Hallmark COMPLEMENT genes with any significant-up call, ranked by number of significant cells; columns are the 12 condition × anatomical-compartment panels. The unit of analysis is one **glomerulus × compartment**, the readout is that unit’s mean log normalized expression and colour is the OLS slope on pseudotime **per 10 pseudotime units** on one symmetric scale (±1.07). Inference is the based on wild cluster bootstrapping over donors (1,999 draws) with BH-FDR **within** each axis × condition × compartment × program × gene-set stratum; **✱ = FDR < 0.05**. **Grey = not testable** — the gene failed the adequacy gate in that panel (split-half Spearman–Brown reliability of the donor mean ≥ 0.50, detected in ≥ 5 donors, ≤ 50 % of its counts in one donor); 55 of the 312 drawn cells are grey, 41 of them in control.

We therefore examined the temporal relationship between complement and another major inflammatory program, IFN-γ, along the injury trajectories, which may represent upstream stimuli for complement production as well as signals downstream complement activation. Complement and IFN-γ response showed closely overlapping dynamics along Lineages 1, 2, and 4, without evidence for a consistent increase in IFN-γ signaling before complement activation **(Sup. Fig COMP1)**. We also observe an increase in spatial autocorrelation of both complement and IFN-γ in the glomeruli, together suggesting a coordinated, but independent engagement as glomerular injury progressed **(Sup. Fig. COMP2)**.

To understand which components of these pathways may be contributing to these signals, we decomposed the complement signal to its constituent genes. We find that genes associated with classical (C1R, C1S, C3), alternative (CFB), and regulatory portions (SERPING1), were associated with higher pseudotime **(Fig. 6d)** and histology based injury, with the strongest signal from LN, followed by FSGS, and then MN in the distal regions of the tissue **(Fig. 6c)**.

### Evidence for complement pathway activation in the tissue surrounding diseased glomeruli transcribing complement pathway components

We extended our analysis to the surrounding periglomerular zone focused on proximal tubule cell morphology and transcriptome **(Fig. 7)**. Using KPMP cell-state labels as a reference for our cohort state assignments, we detected Adaptive and Failed Repair most prominently in FSGS and MN biopsies, while Degenerative dominated control and LN biopsies **(Fig. 7a)**. Interestingly, FSGS biopsies had half as many Visium spots detected as PT compared to control, MN and LN **(Sup. Fig. PTa)**. For each patient, the PT injury fraction was the fraction of PT spots assigned to the Adaptive or Failed Repair states **(Fig. 7b)**. Several variables derived from these state assignments correlated with eGFR, including our PT injury fraction metric (ρ=-0.52) and, most notably, Adaptive PT proportion (ρ=-0.67) **(Fig. 7c)**. Stratified by condition, the injury-fraction–eGFR correlation did not reach significance within any single condition, but eGFR declined with increasing PT injury fraction across all conditions **(Fig. 7d)**. Additionally, we observed enrichment across several complement pathways in the PT spots in our disease conditions, placing complement activity not only in the glomerular structures, but directly within the injured PT compartments, as well **(Fig. 7e**).

**Figure 7.**
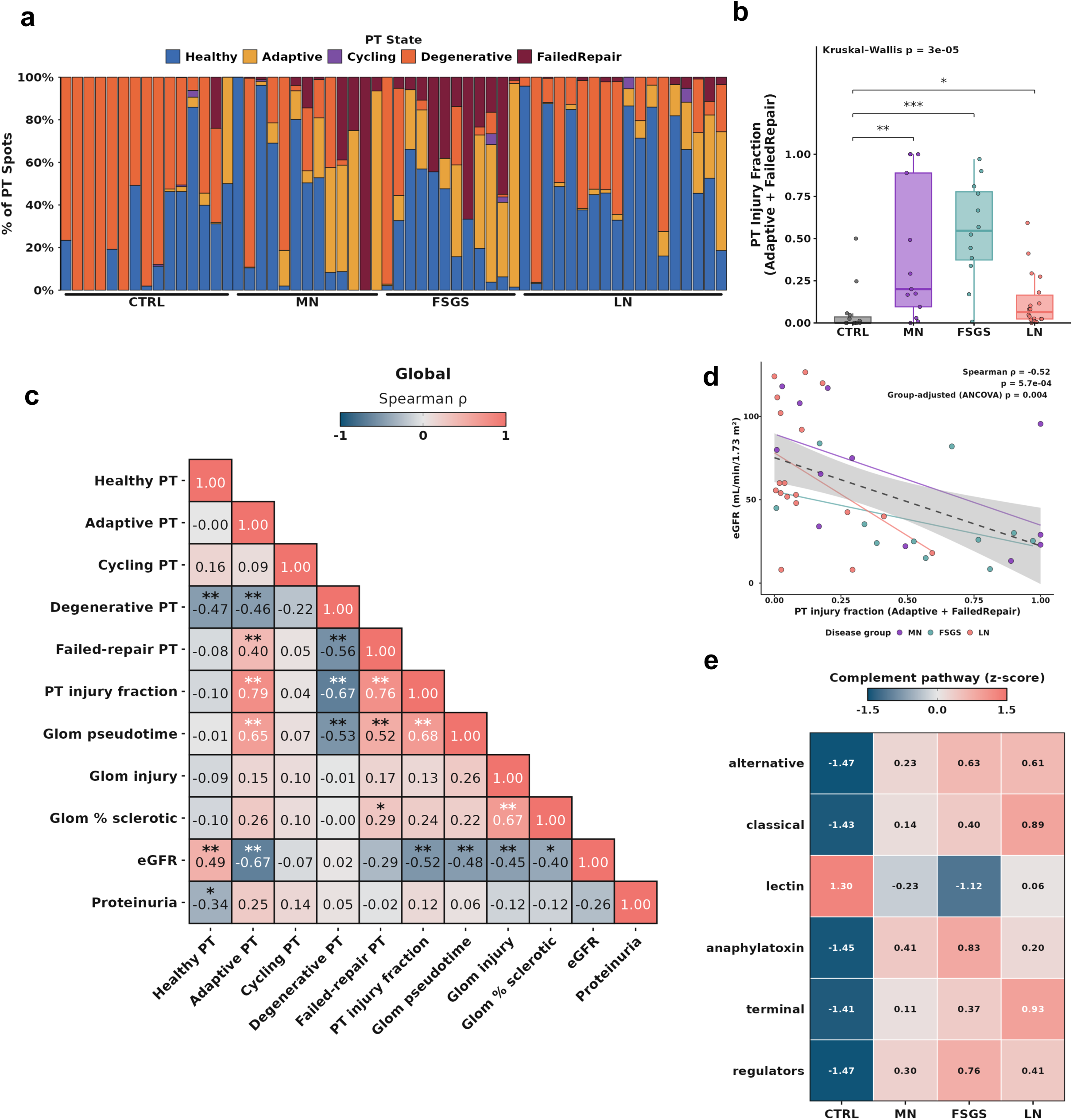
Proximal tubule injury states, kidney function, and complement pathway activity across disease groups. **(a)** Per-patient PT spot composition across five injury states, ordered by increasing PT injury (Adaptive + Failed Repair fraction) within each disease group. **(b)** Per-patient PT injury fraction across the four disease groups (dot = one patient); Kruskal-Wallis test with Bonferroni-corrected comparisons vs. Control. **(c)** Spearman correlation heatmap (all patients) of PT injury states, PT injury fraction, glom pseudotime, glom injury (blinded H&E grade, 1–4), glom % sclerotic (nested-mean fraction graded S2/S3), eGFR, and proteinuria. Cells show rho (color/value) with significance (* p < 0.05, ** p < 0.01); correlations require ≥4 complete pairs, insufficient-data cells greyed. **(d)** PT injury fraction vs. eGFR (n = 41 disease patients; controls excluded, no at-biopsy eGFR). Dot = one patient by disease group; dashed line = global regression, faded lines = per-disease regressions. Reported: global Spearman rho/p and group-adjusted ANCOVA p. Per-group Spearman correlations are shown for each disease group: MN (purple, n = 12, p = 0.060), FSGS (turquoise, n = 11, p = 0.214), LN (coral, n = 18, p = 0.051). **(e)** Complement pathway activity (alternative, classical, lectin, anaphylatoxin, terminal, regulators) in PT spots; each gene set scored per spot (Seurat AddModuleScore, SCT assay), averaged per disease group, shown as z-scores.

### Experimental membranous nephropathy recapitulates the spatial complement program and resolves its cellular sources

Finally, we asked whether the complement-centered program identified in human disease could be reproduced and spatially resolved in an experimental system. We analyzed kidneys from rats with passive Heymann nephritis (PHN), an experimental model of membranous nephropathy, and saline-treated controls at day 14 after serum injection, using complementary molecular approaches, including Visium HD spatial transcriptomics and single-nucleus RNA sequencing **(Fig. 8a; Sup. Table 3)**. The higher spatial resolution of Visium HD enabled analysis not only of the glomerular tuft but also of concentric peri-glomerular regions extending up to 200 μm from the glomerular boundary **(Fig. 8b)**.

**Figure 8.**
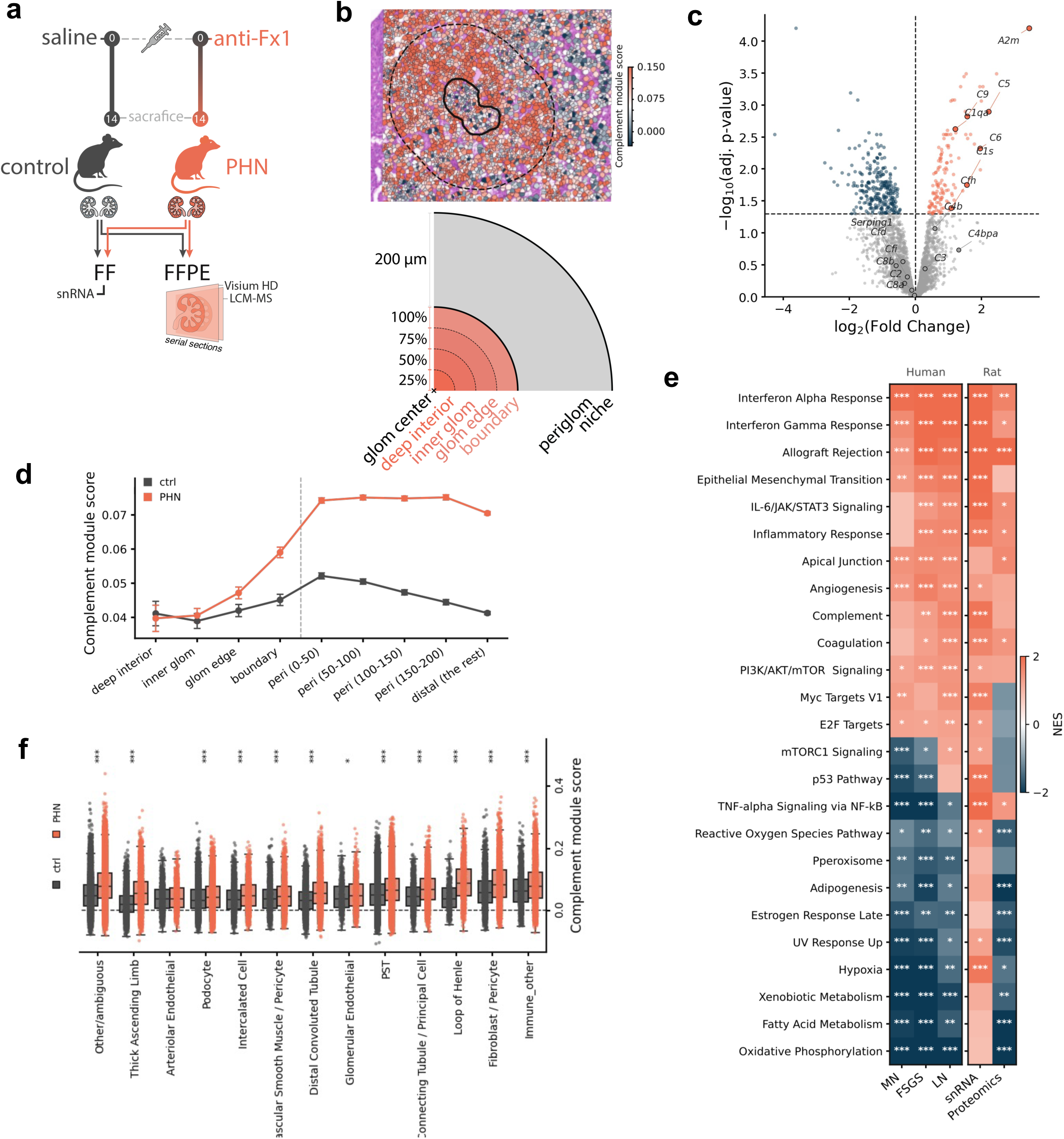
Cross species comparison of membranous nephropathy rat model with passive Heymann nephritis (PHN). (**a**) Experimental design, (**b**) definition of glomerular and peri-glomerular niches, (**c**) volcano plot of differential protein expression analysis highlight complement pathway components, (**d**) Complement module score from glomerular center to distal region (orange: PHN, ctrl: gray) (**e**) GSEA comparison cross-species and within human, across disease groups, (**f**) celltype complement activation comparison indicates activity across multiple cell types.

Topologically, complement pathway activity was increased in PHN relative to control tissue within the glomerulus and extended into the surrounding peri-glomerular niche **(Fig. 8d)**. Thus, complement-associated transcription was not confined to the glomerular tuft but formed a broader spatial field around injured glomeruli. Finally, cross-species pathway comparison demonstrated that major transcriptional programs observed in human glomerular disease were evident in PHN **(Fig. 8e)**. Complement pathway was evident across cell types except arteriolar endothelial cells (Fig. 8f). We also performed mass spectrometry on laser micro-dissected glomeruli in the PHN rats to test the presence of complement components. We observe enrichment of critical proteins in the complement cascade, (C1qa, C6, A2m, C5, Cfh, C9, C1a, C4bpa) **(Fig. 8c)**, confirming that complement pathway activity observed in the glomeruli is detected in the proteome, too.

## DISCUSSION

Here, we integrated histology-guided spatial transcriptomics with trajectory inference and reference-based cell-state deconvolution to define glomerular injury across MN, FSGS, and LN. Despite their distinct initiating mechanisms, these diseases converged on shared transcriptional programs characterized by inflammatory and interferon signaling, complement activation, loss of metabolic homeostasis, and progressive remodeling of glomerular cellular states. At the level of individual glomeruli, trajectory analysis further revealed that these changes were organized along overlapping but disease-biased injury *continua* rather than segregating strictly by diagnosis. Importantly, higher biopsy-level glomerular pseudotime was associated with lower eGFR, linking the spatially resolved molecular state of the glomerulus to kidney function.

Complement emerged repeatedly across independent analytical approaches and was subsequently reproduced in experimental membranous nephropathy, where higher-resolution spatial profiling localized complement-associated transcription to multiple renal cell populations within and surrounding injured glomeruli. Together, these findings support a model in which distinct glomerular diseases retain disease-specific molecular features while converging on a more restricted set of downstream tissue-injury programs. These observations extend prior single-glomerulus spatial profiling in FSGS, MN, and Alport syndrome, which demonstrated marked molecular heterogeneity even among histologically similar glomeruli (Clair, Soloyan et al. 2024), by combining histology-guided glomerular capture with trajectory inference, clinical correlation, and cross-species experimental validation. A central aspect of this study was the integration of transcriptomic information with direct histologic identification of individual glomeruli. Spatial transcriptomics preserves tissue architecture, but the ability to identify a structure transcriptionally remains dependent on preservation of the molecular features used to define that structure. This becomes particularly problematic in advanced disease, where podocyte depletion, dedifferentiation, and sclerosis progressively erode canonical glomerular transcriptional signatures. Consistent with this concept, transcriptomic clustering both included spots outside histologically defined glomeruli and failed to recover a subset of true glomeruli, with the latter discrepancy increasing as histologic injury became more severe. The effect was particularly evident in FSGS, in which sclerotic glomeruli were disproportionately missed by transcriptional classification alone. Histology-guided annotation therefore did more than improve anatomical precision; it prevented systematic exclusion of some of the most severely remodeled glomeruli from molecular analysis.

The transcriptional changes associated with histologic injury reinforced this point. Increasing injury was accompanied by loss of podocyte and filtration-barrier genes, including NPHS2, PODXL, PTPRO, and DDN, together with reduced EHD3, while immune-related transcripts, C1S, and matrix-associated genes increased. These observations suggest that progression is characterized not simply by amplification of disease-specific signaling, but by erosion of differentiated glomerular cell states accompanied by immune activation and extracellular-matrix remodeling. The much larger number of injury-associated genes identified in FSGS and LN than in MN may reflect differences in the degree and nature of structural remodeling represented in the cohort, but it also emphasizes that histologically comparable injury can arise in transcriptionally distinct contexts.

One of the principal findings of the study is that etiologically distinct glomerular diseases converge on common molecular responses. Across MN, FSGS, and LN, glomerular transcriptomes showed concordant enrichment of interferon-associated inflammatory pathways together with suppression of oxidative phosphorylation and fatty-acid metabolic programs. These shared features coexisted with disease-preferential signatures, including TGF-β-associated programs in MN, prominent inflammatory and IL- 6/JAK/STAT3 signaling in FSGS, and strong inflammatory and complement-associated programs in LN (Li, Birmingham et al. 2021, Latt, Heymann et al. 2022, Clair, Soloyan et al. 2024, Hu, Ji et al. 2026). Thus, convergence did not erase disease identity. Rather, the data support an architecture in which diagnosis-specific initiating mechanisms feed into a partially shared downstream response involving inflammation, metabolic dysfunction, and tissue remodeling. These findings are also consistent with single-cell kidney atlases that identify recurrent injury-associated cellular states and niches across etiologically diverse kidney diseases (Lake, Menon et al. 2023).

Trajectory analysis extended this framework beyond static comparisons between diagnostic groups. Individual glomeruli did not occupy discrete disease-specific transcriptional states but were distributed across five inferred trajectories, with control glomeruli enriched in relatively preserved lineages and disease-associated glomeruli preferentially occupying others. Although FSGS, LN, and MN showed different point estimates of representation across Lineages 1, 2, and 4, respectively, these associations were not absolute, emphasizing that the inferred trajectories reflected shared states of injury rather than simple molecular surrogates of diagnosis. This interpretation was independently supported by histology: glomeruli with more severe pathologist-assigned injury accumulated toward later trajectory states.

The molecular organization of these trajectories further supported a common injury continuum. Disease-associated trajectories showed increasing complement, interferon, allograft-rejection, and remodeling programs, whereas another prominent trajectory was characterized by progressive loss of podocyte structural genes together with oxidative phosphorylation, electron-transport, cytoskeletal, and survival programs. Of particular relevance, glomeruli at different pseudotime states frequently coexisted within the same biopsy. This intrabiopsy heterogeneity illustrates a limitation of analyses in which an entire kidney biopsy is treated as a single molecular unit: a sample classified under one diagnosis may simultaneously contain relatively preserved glomeruli and glomeruli that have progressed substantially along an injury trajectory. The inverse association between mean glomerular pseudotime and eGFR further indicates that the distribution of these states is clinically relevant, although the cross-sectional design does not establish that pseudotime represents a true longitudinal sequence within individual glomeruli.

Cell-state deconvolution provided a complementary view of this continuum. Predicted podocyte abundance closely localized to histologically annotated glomeruli and was reduced most prominently in FSGS and LN, whereas MN showed relative preservation.

These differences are consistent with distinct combinations of altered cellular composition and altered cellular state across the three diseases. More broadly, correlations between pseudotime and predicted cell abundance suggested that progression through the glomerular injury landscape is accompanied by coordinated loss of differentiated glomerular populations and expansion of stromal and immune-associated states. Because the Visium platform measures mixtures of cells within each capture spot, these analyses cannot definitively assign individual transcripts to specific cells; nonetheless, the concordance between deconvolution, histology, and trajectory analysis supports the biological relevance of the inferred cellular changes.

Complement was notable because it emerged independently from several levels of analysis. C1S increased with histologic injury, complement-related pathways were enriched in disease-associated pseudotime programs, and complement was particularly prominent in cross-sectional analyses of LN. These observations are important because complement is conventionally considered within the disease-specific biology of individual glomerulopathies. This cross-disease pattern supports complement as an active amplifier of podocyte injury rather than merely a marker of immune-complex deposition. In FSGS, complement activation and IgM/C3 deposition associate with proteinuria and disease severity (Thurman, Wong et al. 2015, Zhang, Gu et al. 2016, Huang, Cui et al. 2020). Our prior work showed that loss of podocyte DAF/CD55 promotes local C3aR–IL-1β signaling and podocyte injury, with similar DAF loss, C3d deposition, and urinary C3a in human FSGS (Angeletti, Cantarelli et al. 2020). More recent data implicate C5a/C5aR1 signaling in FSGS (Gong, Huang et al. 2025), while LN studies link intrarenal complement expression to impaired kidney function and interferon signaling (Tampe, Hakroush et al. 2022). In our present studies, the close pseudotime dynamics of complement and IFN-γ programs did not provide evidence that IFN-γ activation consistently preceded complement, arguing against a simple unidirectional sequence in the present dataset. Rather, these inflammatory modules appeared to become engaged in parallel as glomerular injury advanced. Altogether, our results support testing the efficacy of complement inhibitors in counteracting the progression of chronic kidney diseases (Wooden, Tarragon et al. 2023).

The experimental PHN studies extended these human observations in two ways. First, the model recapitulated complement-associated transcriptional changes observed in human disease, supporting conservation of this response across species. Second, Visium HD provided sufficient spatial resolution to interrogate complement activity within the glomerulus and its surrounding niche. Complement-associated transcription was increased not only within injured glomeruli but also across peri-glomerular regions, indicating that the response extends beyond the glomerular tuft. Moreover, increased complement module activity was distributed among multiple renal populations, including podocytes, endothelial cells, fibroblast/pericyte populations, immune cells, and tubular compartments. These data argue against interpreting the complement signature solely as a marker of circulating or infiltrating immune activity and instead support a substantial local tissue contribution to the complement-associated transcriptional environment of glomerular injury.

This spatial organization may have broader implications for how progressive glomerular disease is conceptualized. Conventional diagnostic categories remain essential because they identify initiating mechanisms and guide disease-specific therapy. Our findings suggest a complementary framework in which progression is also defined by the position of individual glomeruli along shared tissue-injury states. In this model, two patients with the same diagnosis could differ substantially in the proportion of glomeruli occupying preserved versus advanced states, whereas patients with different diagnoses could share downstream inflammatory, metabolic, or remodeling programs. Such molecular convergence may help explain why distinct glomerular diseases ultimately develop common structural outcomes such as glomerulosclerosis and progressive loss of filtration. It also raises the possibility that therapies directed at shared downstream processes could complement treatments aimed at the initiating cause of disease. The present study nominates complement as one such convergent pathway, but does not establish that complement activation is itself the principal driver of progression or that complement inhibition would have equivalent effects across these disorders.

Several limitations should be considered. First, conventional Visium capture spots contain multiple cells and may include transcripts from adjacent glomerular, vascular, interstitial, or tubular compartments; cell-type deconvolution therefore remains inferential. The higher-resolution PHN analysis helps address this limitation experimentally but does not substitute for equivalent high-resolution profiling of the human biopsies. Second, the human cohort is relatively modest and includes heterogeneity in disease severity, treatment exposure, tissue quality, and number of glomeruli captured per biopsy. Third, the study is cross-sectional. Pseudotime provides a biologically coherent ordering of transcriptional states but cannot establish that an individual glomerulus will progress through those states longitudinally. Fourth, although mean pseudotime correlated with baseline eGFR, the available data do not establish that the spatial injury trajectories predict subsequent kidney-function decline independently of conventional clinical or histologic variables. Finally, the complement analyses demonstrate transcriptional enrichment and spatial localization but do not directly measure complement cleavage, deposition, or functional activation throughout the human cohort. Spatial RNA abundance also cannot by itself establish protein synthesis, secretion, or pathway flux, or determine how much of the tissue complement protein pool derives from local synthesis versus plasma-derived complement. The PHN glomerular proteomic data provide orthogonal evidence for selected complement components, but equivalent protein-level validation is not available across the human cohort. Orthogonal protein-level and functional studies will therefore be important to determine which components of the complement cascade are active, which renal cell populations contribute most importantly to that activity, and whether complement participates causally in progression.

In summary, histology-guided spatial transcriptomics reveals that MN, FSGS, and LN are organized not only by diagnosis-specific molecular programs but also by shared glomerular injury states. Individual glomeruli occupy heterogeneous positions along overlapping transcriptional trajectories characterized by progressive loss of podocyte and metabolic programs and increasing inflammatory, complement, and remodeling activity. The relationship between pseudotime and kidney function links these spatial states to clinically relevant injury, while cross-species Visium HD analysis identifies a multicellular, locally distributed complement program extending beyond the glomerular tuft. These findings provide a spatial framework for understanding how distinct glomerular diseases converge during progression and identify local complement-associated signaling as a prominent component of that shared injury response.

## METHODS

### Patients

Patient and control biopsies were obtained from paraffin blocks at pathology biorepository at Mount Sinai Hospital, New York, NY and deidentified. The Institutional Review Board approved the protocol for the collection of human samples. Informed consent for kidney biopsies was obtained from participants.

### Clinical outcomes and statistical analysis

Individual estimated glomerular filtration rate (eGFR) slopes were estimated using linear mixed-effects models: eGFR ∼ time after biopsy + (1 + time after biopsy | patient). Patients with at least three eGFR observations were included. Annual eGFR decline was categorized as rapid (<= -5), slow (-5, 0], or improvement (>0) mL min-1 1.73m-2 yr-1. Baseline differences between decline groups were assessed by Kruskal-Wallis testing followed by pairwise, two-sided Mann-Whitney U tests with Bonferroni correction. Unless otherwise specified, inferential analyses used the biopsy, glomerulus, or tissue-region unit indicated, and were two-sided.

#### Tissue sectioning

Formalin-fixed paraffin-embedded (FFPE) kidney biopsies were sectioned at 5-–7 µm using a microtome with RNAse-free practices. Sections were mounted onto 10x Genomics Visium Spatial Gene Expression Slides (FFPE) according to the manufacturer’s instructions and baked to promote adherence. Adjacent sections were mounted on standard glass slides for hematoxylin and eosin (H&E) to support histologic annotation.

#### Deparaffinization and pre-treatment

Slides were deparaffinized and rehydrated using xylene and graded ethanols, followed by antigen retrieval and tissue permeabilization using the Visium FFPE workflow reagents and conditions recommended by 10x Genomics. To enable probe-based transcript capture, tissue sections were equilibrated and processed to optimizeaccessibility while preserving morphology. All steps were performed with appropriate controls to minimize RNA degradation and batch effects.

### Probe hybridization and ligation

Spatially barcoded gene expression was generated using the 10x Genomics Visium Spatial Gene Expression Reagent Kits for FFPE. Briefly, a whole-transcriptome probe panel was hybridized to the tissue section, followed by extension and ligation to generate amplifiable products corresponding to targeted RNA molecules. After ligation, probes were released and collected from each capture area to retain spatial barcodes linked to each spot location.

#### Library preparation and sequencing

Sequencing libraries were constructed following the Visium FFPE protocol, including PCR amplification, indexing, and size selection. Library quality and fragment size distribution were assessed using capillary electrophoresis, and concentrations were quantified by fluorometric methods. Libraries were pooled and sequenced on an Illumina platform with paired end reads using the 10x Genomics recommended read structure (including spatial barcode and UMI reads). Sequencing depth was targeted based on the number of capture areas and tissue quality, consistent with manufacturer guidance.

#### Imaging and spatial alignment

Image analysis was performed using HALO AI software (Indica Lab, LLC – indicalab.com/halo-ai). HALO AI classifier was created using DenseNetV2 by Indica Labs modeling two separate classes of healthy and unhealthy glomeruli and a remainder class of surrounding tissue. Several H&E images from the cohort were randomly selected and used as a training set with three ordinal classes of equal weight: tissue, glomeruli, abnormal glom. Learning rate was set at 0.05 and batch size at 8. Once the training was completed with a final algorithm, all the H&E images went through image analysis. After the completion of the image analysis, all images were visually reviewed to confirm the proper glomeruli detection. The data generated was then used for spatial alignment with transcriptome.

#### Human spatial transcriptomics and glomerular annotation

Human kidney tissue was profiled with Visium spatial transcriptomics, for which the physical capture-spot diameter was 55 µm. A HALO AI classifier was applied to H&E-stained sections to identify glomeruli, and all images were subsequently visually reviewed. Counts were normalized to 10,000 per spot and log1p-transformed. Genes used for clustering were detected in at least five spots in one condition and in at least 10 spots overall. We selected 2,000 highly variable genes, calculated 50 principal components, and performed Harmony integration by manually assigned region. A 15- nearest-neighbor graph was used for UMAP embedding and Leiden clustering (seed 42). The glomerular transcriptional population was defined as Leiden cluster 2 and supported by expression of the podocyte markers *NPHS1*, *NPHS2*, and *PODXL*.

#### Histologic annotation and integration

Glomeruli were scored using standard clinical grading systems: the Oxford Classification S-score and the Glomerulosclerosis Index (GSI) (Trimarchi, Barratt et al. 2017, Wang, Wang et al. 2021). For each biopsy, individual glomeruli were cropped from high-resolution histology images and scored independently by a trained histologist who was blinded to patient identity and disease status using a self-developed web-tool.

#### Glomerular pseudobulk differential expression and enrichment

Raw counts from annotated glomerular spots were aggregated by biopsy. Differential expression was performed in DESeq2 using the design ∼ condition, comparing each of membranous nephropathy (MN), focal segmental glomerulosclerosis (FSGS), and lupus nephritis (LN) with control (CTRL). Genes with a total count of at least 10 were tested using the Wald test. Significance was defined by DESeq2-adjusted p < 0.05, without a log-fold-change threshold. Geneset enrichment analysis was performed with gseapy against MSigDB Hallmark 2020.

#### Glomerular trajectory and program analysis

Slingshot was applied to the glomerular embedding with approx_points = min(300, n), thresh = 0.01, stretch = 0.8, allow.breaks = FALSE, and shrink = 0.99. A weighted mean pseudotime was calculated from lineage pseudotimes using curve weights. tradeSeq fitGAM models used six knots, followed by association and differential-end tests; selected genes met BH FDR < 0.05. For trajectory program analysis, fitted smooth curves were z-scored per gene. Dynamic tree cutting used correlation distance, average linkage, deepSplit = 0, and a minimum cluster size of min(30, 0.3*n_genes). Programs were tested with Enrichr against GO Biological Process 2026 and MSigDB Hallmark 2020; clusters containing fewer than five genes were omitted, and adjusted p < 0.05 was required.

#### Spatial processing and PT injury-state assignment

Spatial count matrices were processed in R (Seurat v5). Spots passing per-spot QC, a Lake et al. 2025–style in-tissue edge filter (<6 neighbors), and >200 UMIs were retained (41,581 spots); immune-infiltrate spots from the 50957 Normal control region were excluded. Spots were SCTransform-normalized per slide with PCA on SCT residuals. Cell states were transferred from the KPMP adult kidney snRNA-seq atlas (v2; 1.39M nuclei) by anchor-based label transfer using SubclassLevel3 annotations (128 states). The reference was subsampled proportional to state abundance (∼116,700 nuclei; floor 200/state), SCTransform-normalized, and anchors identified via FindTransferAnchors (SCT, reference PCA, 30 dims); TransferData projected labels onto each spot (recompute.residuals = FALSE). Spots were assigned to compartments using the KPMP "Niche analysis (Visium)" procedure (SubclassLevel3→SubclassLevel1→Class): stage one assigned the majority Class, stage two subdivided epithelial spots by top SubclassLevel1 type (>20%). PT spots were further restricted to those confirmed as tubule by HALO AI/histopathologist annotation (8,889 of 9,998). Per PT spot, SubclassLevel3 proportions were aggregated into five states—Healthy (PT-S1/2/3), Adaptive (aPT), Cycling (cycPT), Degenerative (dPT), Failed Repair (frPT)—and labeled by argmax. PT niches were also defined by Louvain clustering (SNN graph, k = 20) of proportion vectors.

#### PT injury quantification and clinical correlation

Per patient, PT injury fraction was the proportion of Adaptive + Failed-Repair spots, and degenerative fraction the proportion of Degenerative spots, among that patient’s PT spots. Associations with eGFR (n = 41) and proteinuria (n = 38; log10) were assessed by Spearman correlation (across and within groups) plus group-adjusted ANCOVA (outcome ∼ predictor + group); controls were excluded. Group comparisons used Wilcoxon rank-sum (two groups) or Kruskal-Wallis with Bonferroni-corrected comparisons vs. Control (multiple groups), with Benjamini-Hochberg correction unless stated. Analyses used R (Seurat v5, edgeR, clusterProfiler/fgsea, patchwork, ggplot2) and Python (scanpy).

### Rat cross-species validation

Passive Heymann Nephritis (PHN) was induced in male RjHan:SD rats (Janvier; housed two per cage under a 12:12 h light/dark cycle) by a single intravenous (tail vein) injection of sheep anti-Fx1A serum (Probetex, Lot #607-4T; 4 mL/kg) on Day 0, under license BS- 2917. At necropsy (Day 15), kidneys were collected and one portion was snap-frozen for downstream molecular analyses. Nuclei were isolated from snap-frozen kidney tissue (≤50 mg) using the Chromium Nuclei Isolation Kit (10x Genomics, PN-1000494) and processed with the Chromium Next GEM Single Cell 3′ Reagent Kit v3.1 (Dual Index; 10x Genomics, protocol CG000315). cDNA and final libraries were quality-controlled by Bioanalyzer/TapeStation (High Sensitivity DNA assay) prior to sequencing on an Illumina NovaSeq 6000 platform (paired-end, dual-index; R1 28 cycles, I7/I5 10 cycles each, R2 90 cycles). Following quality control, single-nucleus RNA-seq data from five vehicle- treated PHN rats and three healthy control rats were retained for downstream analysis of glomerular and podocyte cell populations.

For spatial transcriptomics and proteomic profiling of glomeruli, consecutive 5 µm sections were cut from the same FFPE kidney blocks, with one section processed for Visium HD spatial gene expression and the adjacent section used for laser-capture microdissection coupled to mass spectrometry (LCM-MS). Sections were dried, H&E stained and imaged, then destained and decrosslinked, followed by probe hybridization and library preparation according to the 10x Genomics Visium HD FFPE Tissue Preparation Handbook (CG000684, Rev D) and Visium HD Spatial Gene Expression Reagent Kit User Guide (CG000685, Rev D). Sequencing libraries were generated using 10 PCR cycles for sample indexing, quality-controlled on a TapeStation 4200 (D1000 assay) and Qubit 4.0 (dsDNA HS assay), and pooled according to capture area prior to sequencing on a NovaSeq X+ (1.5B flow cell, 320 pM loading; run 20260121_LH00759_0090_B22GGLFLT1). Following TapeStation-based quality assessment, four sections (2 PHN vehicle-treated, 2 healthy) were selected for downstream processing. Raw sequencing data were processed with Space Ranger (v4.0.1; rat reference genome 2.1.0). For LCM-MS, glomeruli were microdissected from the section adjacent to each Visium HD section, pooled per animal, and analyzed by mass spectrometry-based proteomics.

snRNA-seq nuclei were QC-filtered on genes per cell (100 - 8000), ≤30%% mitochondrial reads, and ≤50,000 total counts, with genes retained if detected in ≥3 cells. Cell type labels were transferred from a GSE209821 rat kidney reference genes were intersected between query and reference, 2000 HVGs selected on the reference, a 50-PC reference PCA/neighbor/UMAP embedding built, and sc.tl.ingest used to project query nuclei into that space and assign each the reference label of its neighbors (Balzer, Pavkovic et al. 2023).

Podocytes pseudobulk was compared with DESeq2 using the design ∼ condition. Genes with total count >= 10 were tested, with adjusted p < 0.05 defining significance. Cross-species comparisons were restricted to one-to-one human-rat orthologs present in the rat expression matrix. Complement module scoring used the same approach as in human tissue.

In Visium HD data, glomeruli were segmented using HALO AI as described above. We used the segmented cell output of Space Ranger for analysis. Cell type deconvolution of the 8 µm Visium HD bins was performed with cell2location: a negative-binomial regression model was first trained on the GSE209821 snRNA-seq kidney reference (clusters2 labels, batch key orig.ident, 250 epochs) to estimate per-cell-type expression signatures, and the spatial model was then fitted per sample over the gene intersection across all samples (full-batch, up to 30,000 epochs, N_cells_per_location=3, detection_alpha=20, in 50,000-bin chunks). The celltype was derived as the argmax of the cell2location probabilities. Peri-glomerular tissue was defined as 0-200 µm from the glomerular boundary and distal tissue as >200 µm. Complement-associated mRNA programs were evaluated using the MSigDB Hallmark COMPLEMENT gene set and the ssGSEA with a minimum gene-set size of 5, a maximum size of 1,000, and seed 0.

For the mass spectrometry, precursor-level intensities from the Spectronaut DIA were normalized using median-of-ratios scaling. Protein abundances were computed from the filtered, normalized precursor data via MaxLFQ (q≤0.01), yielding the final protein-level abundance table. Differential protein abundance was assessed with limma in R using a no-intercept design with Healthy and PHN group columns. The PHN minus Healthy contrast was tested with lmFit, contrasts.fit, and empirical-Bayes moderation using eBayes. Proteins with adjusted p < 0.05 were considered significant.

## Data Availability

All data produced in the present study are available upon reasonable request to the authors

## ACKNOWLEDGEMENTS

This research was supported by the Novartis Postdoctoral Fellowship Program and the Novartis Post-Baccaloreate Program, Biomedical Education and Innovation, Novartis Biomedical Research, Novartis Pharma AG. The content is solely the responsibility of the authors. Additionally, we thank Markus Stoeckli and Bill Dietrich for their leadership during this collaboration, and we thank Miguel Fribourg-Casajuana for the thoughtful discussions.

## AUTHOR CONTRIBUTIONS

P.C., E.O., M.M., F.T., V.D. contributed to the conception and design of the work. P.C., J.L., T.S., S.W., P.C., J.H.-G. contributed to the analysis and interpretation of the data, C.O., S.B., S.W, J.J.-W., J.P. provided contributed to data generation. X.W., K.M., T.S., S.W., P.C., J.H.-G., J.L., P.C. drafted and revised the work.

## DATA AVAILABILITY

The code for spatial and transcript data is available for review as a set of available reports provided as accessory material. Data are available for review and in process of being deposited into public repository.

---

**Sup. Fig. CLINIC.**
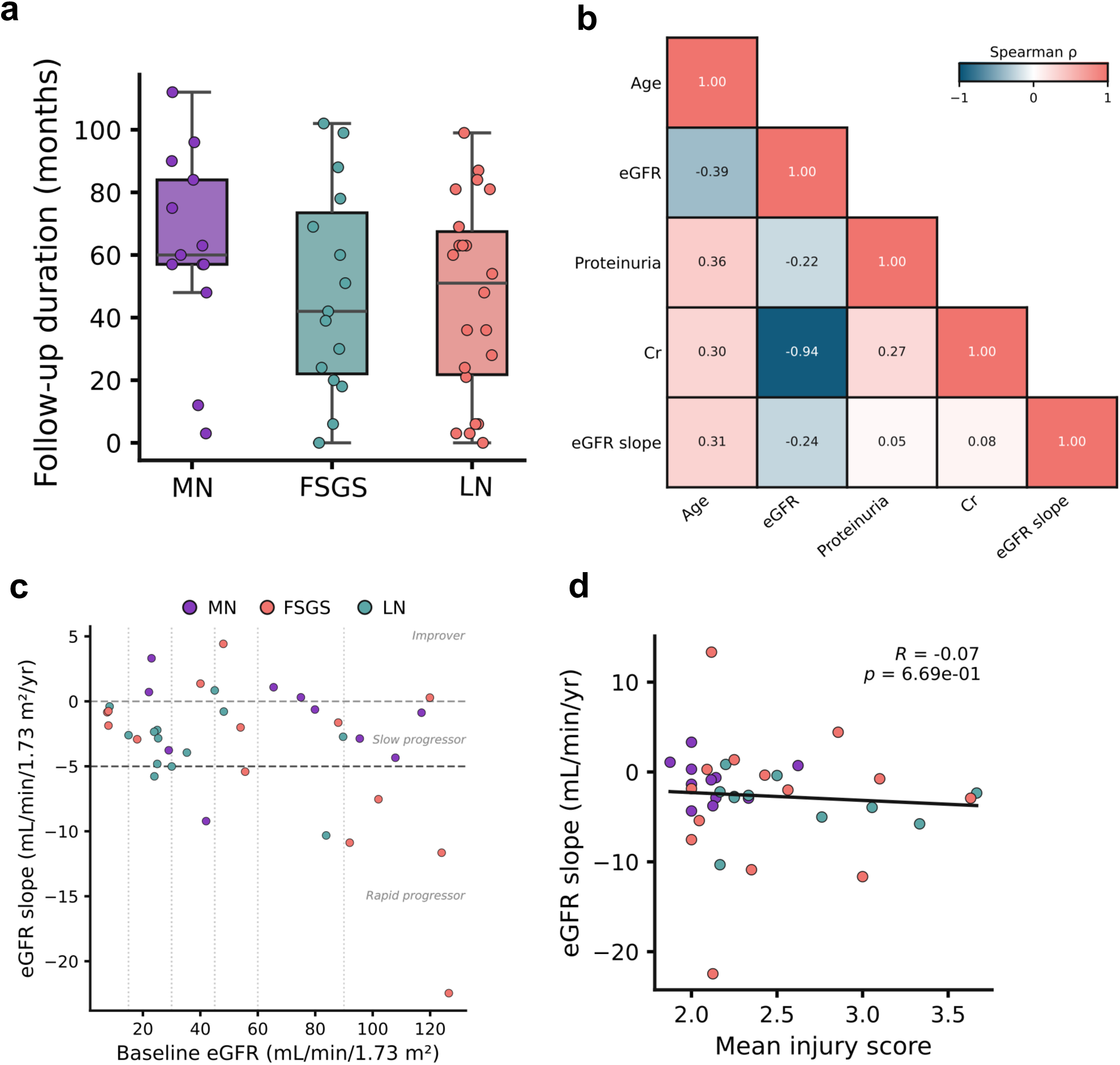
Clinical characteristics and longitudinal kidney function of the study cohort. a) Follow-up duration of patients in months. b) Correlation heatmap of clinical variables across disease. c) Annual eGFR slope versus baseline eGFR, colored by disease group. Horizontal dashed lines mark progressor class thresholds (Improver: slope > 0; Slow progressor: −5 to 0; Rapid progressor: < −5 mL/min/1.73 m²/yr); vertical dotted lines mark CKD stage boundaries by baseline eGFR. Comparisons to CTRL (Dunn’s test for continuous; pairwise Fisher’s exact for categorical), with Holm correction across the three comparisons. Kidney health clinical values are unavailable for all controls

**Sup. Fig. PSEUDOTIME.**
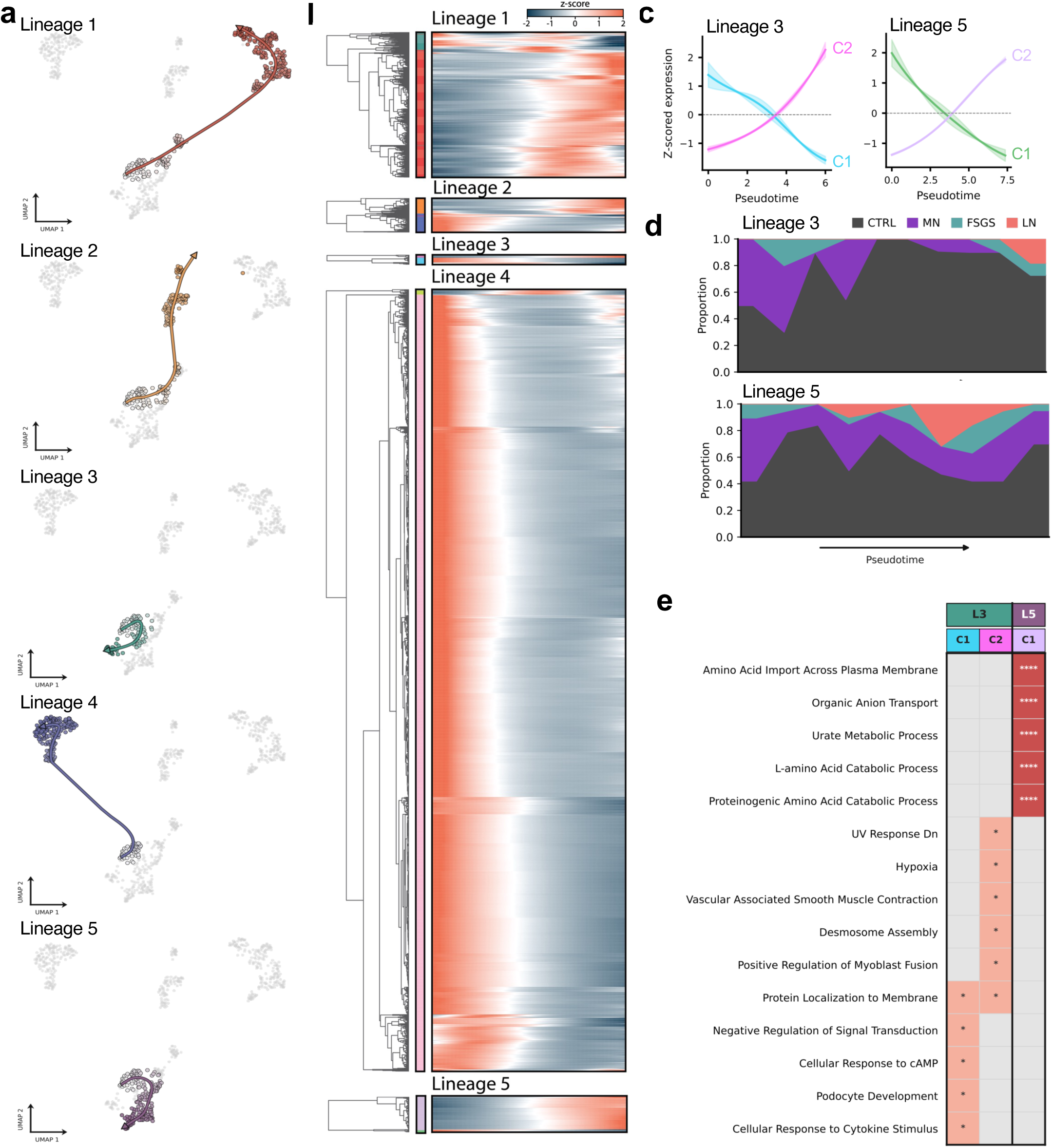
Pseudotime trajectory. **(a)** UMAP of pseudobulked glomerular transcriptomes, with each of the five Slingshot lineages individually highlighted (colored) against all glomeruli (grey). **(b)** Heatmap of z-scored expression along pseudotime for genes significantly associated with each lineage, with hierarchical clustering dendrogram (row color bar indicates cluster assignment). **(c)** Hierarchical clustering of significantly pseudotime-associated genes, shown as smoothed z-scored expression per cluster. **(d)** Disease composition along pseudotime for Lineage 3 and Lineage 5. **(e)** Top 5 enriched pathways per gene cluster (C1–C2) within each lineage (L3, L5).

**Sup. Fig. PSEUDOTIME2.**
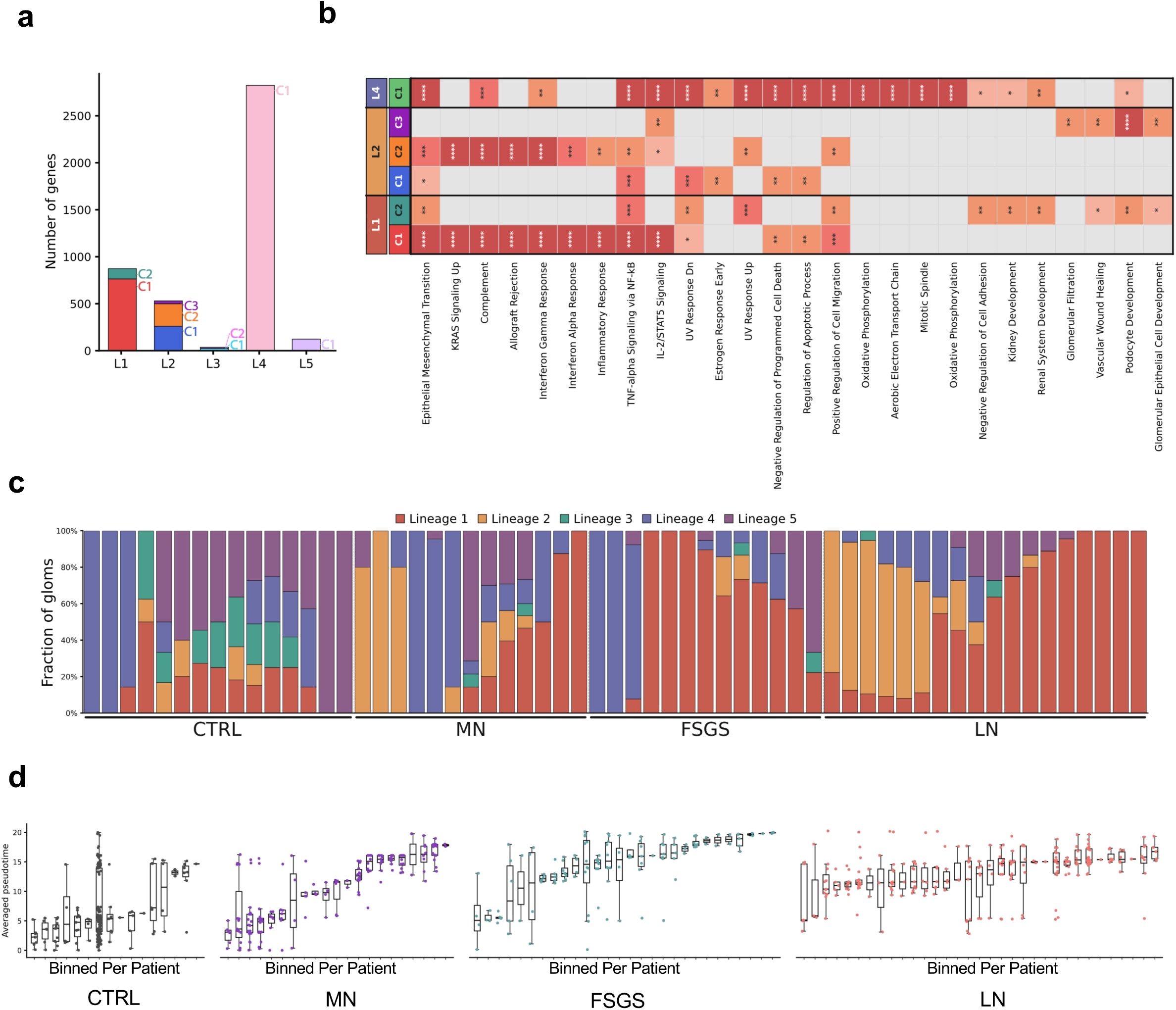
(a) Number of genes associated with each pseudotime lineage. **(b)** Pathway enrichments associated to lineages **(c)** per-patient lineage commitment. **(d)** per patient glom pseudotime variability stratified by disease.

**Sup. Fig. CELLDECONVOLVE.**
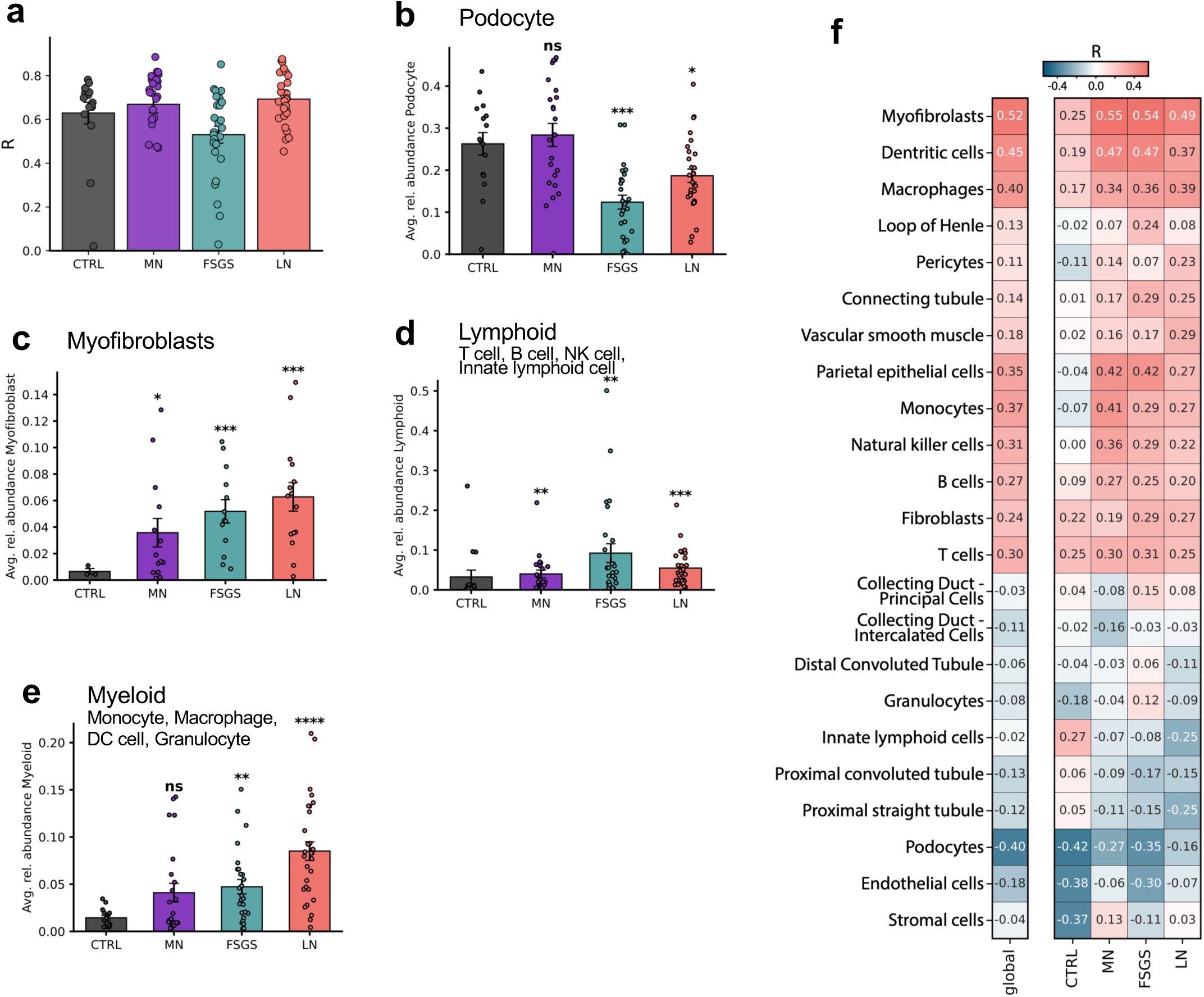
Spatial cell state changes in glomerular injury across disease conditions **(a)** Point–biserial correlation (R) between visually identified glomerular spots (binary) and podocyte abundance scores per condition; bars show mean ± s.e.m. across samples and dots indicate individual samples. **(b)** podocyte detection is reduced in FSGS as compared to the other groups, **(c)** Myofibroblasts are elevated in FSGS and LN, **(d)** lymphoid cells are elevated in all disease groups, **(e)** myloid cells are elevated in all disease groups. **(f)** Global and per disease state correlations of cell types.

**Sup. Fig. COMP1.**
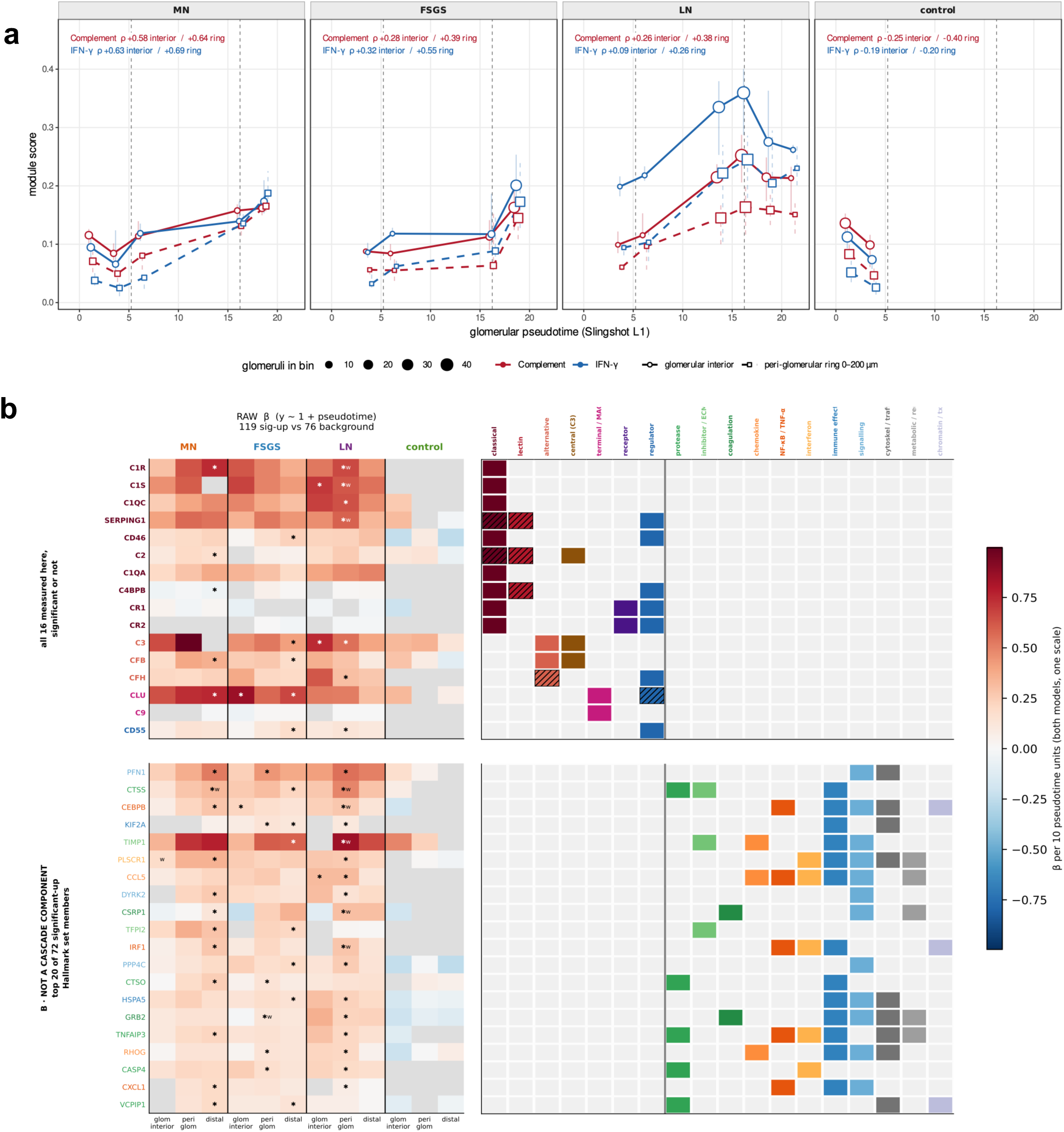
(a) Per-glomerulus Hallmark module scores for COMPLEMENT (red) and INTERFERON GAMMA RESPONSE (blue) plotted against glomerular pseudotime (Slingshot lineage L1), separately for membranous nephropathy (MN), focal segmental glomerulosclerosis (FSGS), lupus nephritis (LN) and non-diseased control kidney. **The unit of analysis is one glomerulus** (*\*n\** = 280 glomeruli from 46 donors: MN 51/9, FSGS 60/11, LN 116/16, control 53/10), restricted to glomeruli with an estimable L1 pseudotime (lineage weight *\*w\** L1 > 0). ρ printed in each panel is **Spearman’s correlation computed on the individual glomeruli**, not on the plotted medians, given separately for the interior and the ring. **(b)** Heatmap of complement pathway members (top) and top remaining genes (bottom) faceted by disease type and distance from glom center. Adjacent is a pathway/process annotation for listed genes.

**Sup. Fig. COMP2.**
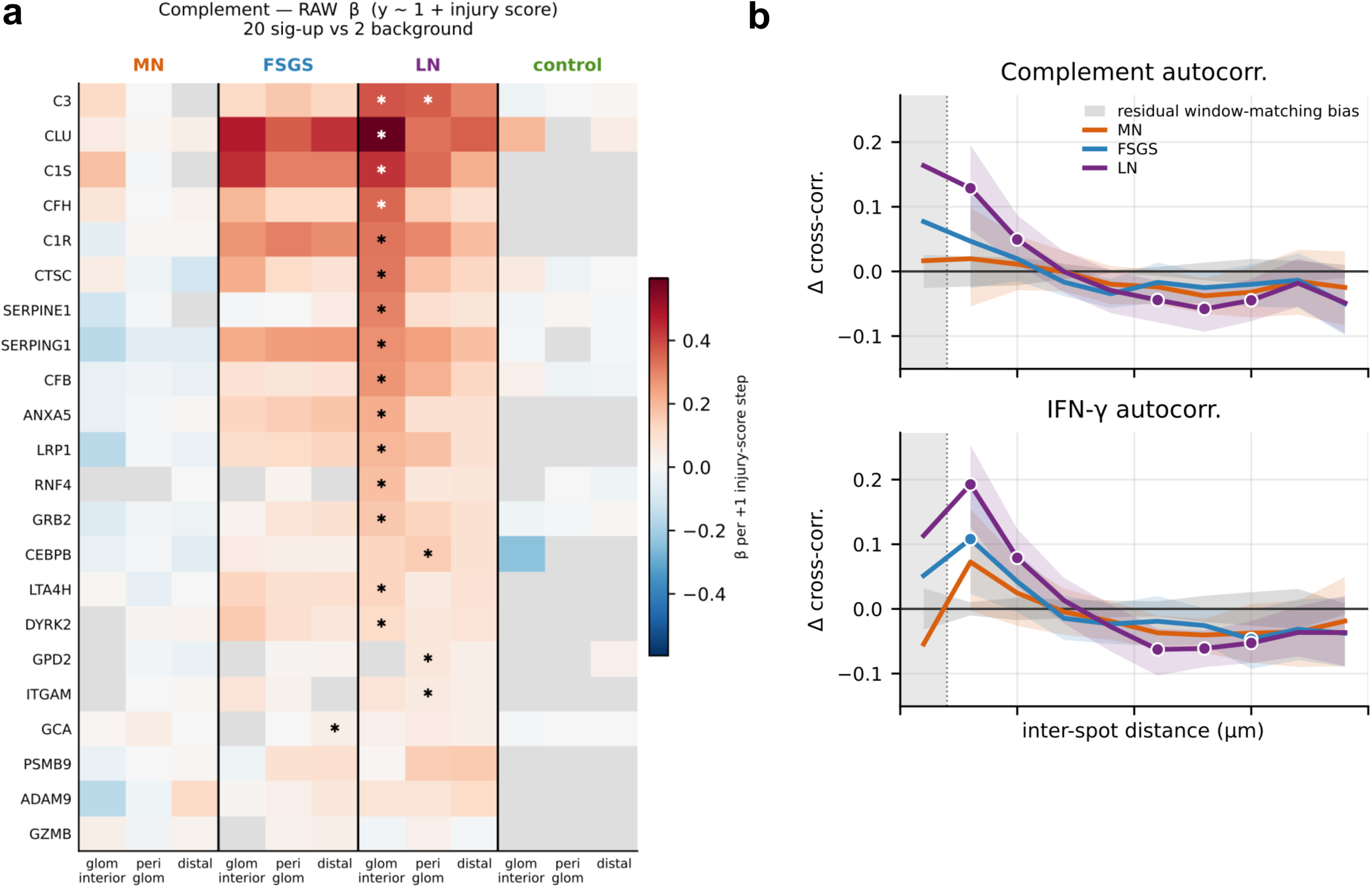
(a) Per-gene β of Hallmark complement genes vs the pathologist injury score (1–4), raw model, per glomerulus × compartment unit, donor-clustered; ✱ = FDR<0.05, grey = not testable. 20 significant-up cells vs 2 background — 19 of them LN, none in MN or control, on an axis with only 4 distinct values (60–87 % ties), so the MN/control zeros are untested, not negative. **(b)** Complement pathway (top) and INF- gamma (bottom) autocorrelation plots .

**Sup. Fig. PT.**
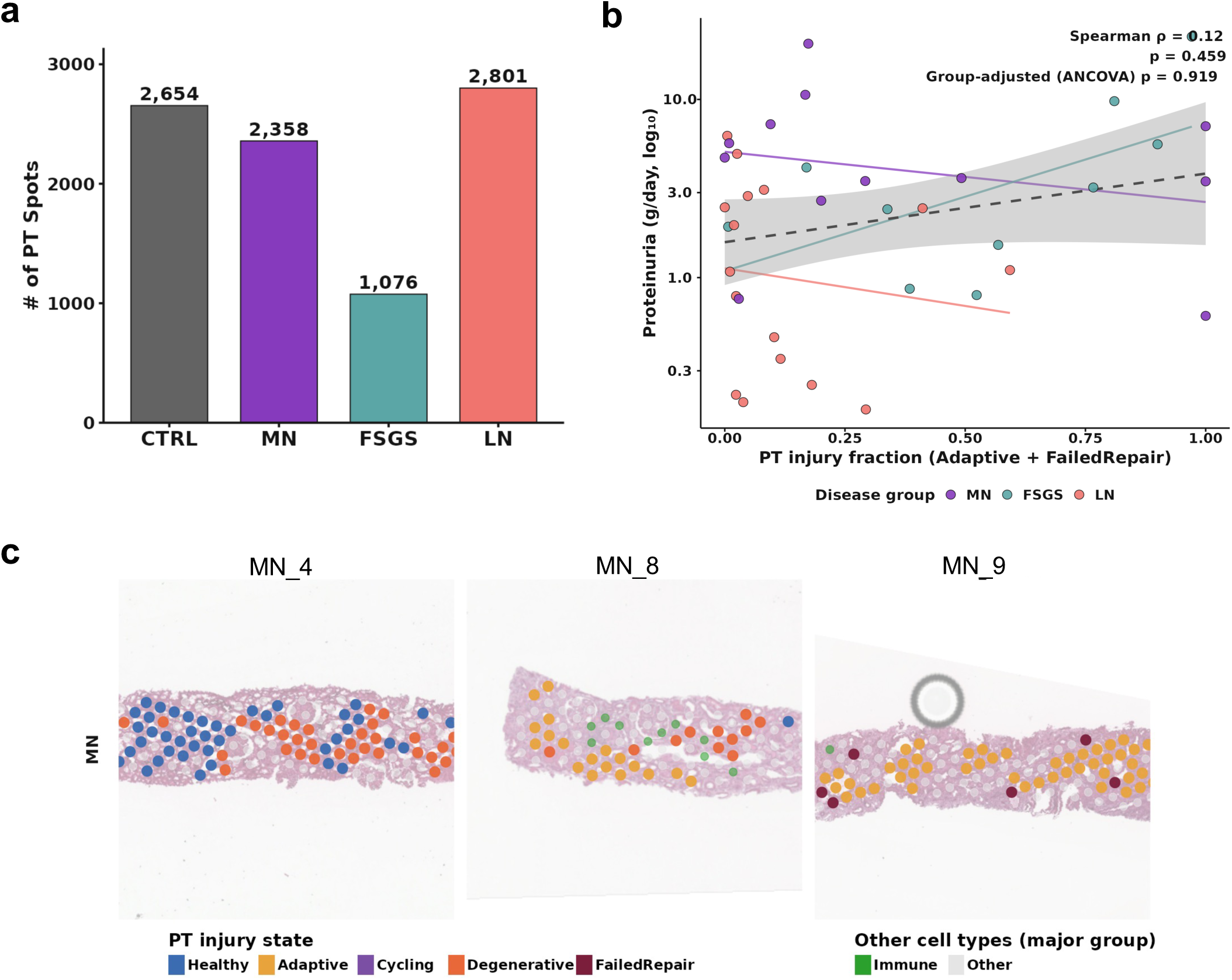
- Proximal tubule injury states in relation to proteinuria and tissue architecture. **(a)** Number of PT spots detected by label transfer and confirmed as tubule per condition. **(b)** PT injury fraction vs. proteinuria (g/day, log10 scale; n = 38 disease patients; controls excluded, no at-biopsy proteinuria). Dot = one patient by disease group; dashed line = global regression, faded lines = per-disease regressions. Reported: global Spearman rho/p and group-adjusted ANCOVA p. Per-group significance marked by colored asterisk: MN (purple, n = 12, p = 0.351), FSGS (turquoise, n = 10, p = 0.088), LN (coral, n = 16, p = 0.165 → 0.213). **(c)** Representative H&E Visium sections with spots colored by PT injury state (Healthy, Adaptive, Cycling, Degenerative, Failed Repair) or major cell group for non-PT spots (TAL, GLOM, EC, Stromal, CD, DCT/CNT, DTL). Three samples represent the minimum, median, and maximum PT injury fraction. States were assigned via Seurat label transfer from the KPMP snRNA-seq reference (SubclassLevel3), followed by Lake et al. 2025 major- group classification.

**Sup. Table 1.** Baseline characteristics by disease group. Patients are grouped as control (CTRL), membranous nephropathy (MN),focal segmental glomerulosclerosis (FSGS), and lupus nephritis (LN). Continuous variables are shown as mean (SD); categorical variables as n (%). Asterisks denote significant differences versus the CTRL reference group: *p<0.05, **p<0.01, ***p<0.001, ****p<0.0001. Significance was assessed hierarchically, an omnibus test (Kruskal–Wallis for continuous, Fisher’s exact for categorical variables) followed, only if significant, by post-hoc comparisons of each disease.

| Variable | Category | CTRL | MN | FSGS | LN |
| --- | --- | --- | --- | --- | --- |
| n |  | 15 | 14 | 15 | 22 |
| Age (years), mean (SD) |  | 61.1 (12.4) | 54.6 (14.8) | 57.3 (13.0) | 35.7 (14.3)**** |
| Sex, n (%) | Female | 6 (40.0) | 5 (35.7) | 8 (53.3) | 19 (86.4)* |
|  | Male | 9 (60.0) | 9 (64.3) | 7 (46.7) | 3 (13.6) |
| Self-reported race, n (%) | Asian | 0 (0.0) | 0 (0.0) | 1 (6.7) | 3 (13.6) |
|  | Black | 0 (0.0) | 3 (21.4) | 7 (46.7) | 8 (36.4) |
|  | Hispanic | 0 (0.0) | 1 (7.1) | 3 (20.0) | 8 (36.4) |
|  | Mixed | 0 (0.0) | 0 (0.0) | 1 (6.7) | 0 (0.0) |
|  | White | 0 (0.0) | 9 (64.3) | 1 (6.7) | 2 (9.1) |
|  | Unknown | 15 (100.0) | 1 (7.1) | 2 (13.3) | 1 (4.5) |
| Self-reported ethnicity, n (%) | Hispanic | 0 (0.0) | 1 (7.1) | 4 (26.7) | 9 (40.9) |
|  | Non-Hispanic | 0 (0.0) | 10 (71.4) | 8 (53.3) | 12 (54.5) |
|  | Unknown | 15 (100.0) | 3 (21.4) | 3 (20.0) | 1 (4.5) |
| eGFR at biopsy (ml/min/1.73m<sup>2</sup>), mean (SD) |  | – | 55.9 (34.7) | 33.7 (17.3) | 53.0 (27.1) |
| Proteinuria at biopsy (g/day), mean (SD) |  | – | 5.9 (5.4) | 4.7 (6.1) | 1.8 (1.8) |
| Creatinine at biopsy (mg/dL), mean (SD) |  | – | 1.7 (1.2) | 2.4 (1.4) | 1.9 (2.0) |
| Annual eGFR slope (ml/min/1.73m<sup>2</sup>/yr), mean (SD) |  | – | 0.7 (2.9) | -3.9 (4.7) | -0.4 (11.1) |
| Follow-up (months), mean (SD) |  | – | 62.6 (30.7) | 48.4 (33.1) | 46.1 (31.4) |
| N eGFR timepoints, mean (SD) |  | – | 11.8 (7.0) | 12.2 (8.6) | 9.9 (7.4) |
| N glomeruli profiled, mean (SD) |  | 13.7 (30.7) | 13.8 (12.0) | 10.2 (4.4) | 13.9 (9.6)* |
| Anti-PLA2R status, n (%) | Positive | 0 (0.0) | 4 (28.6) | 0 (0.0) | 0 (0.0) |
|  | Negative | 0 (0.0) | 8 (57.1) | 0 (0.0) | 0 (0.0) |
|  | Unknown | 15 (100.0) | 2 (14.3) | 15 (100.0) | 22 (100.0) |
| On treatment at biopsy, n (%) | Yes | 0 (0.0) | 0 (0.0) | 0 (0.0) | 2 (9.1) |
|  | No | 0 (0.0) | 14 (100.0) | 15 (100.0) | 20 (90.9) |
|  | Unknown | 15 (100.0) | 0 (0.0) | 0 (0.0) | 0 (0.0) |

**Sup. Table 2.** – Differential expression analysis of pseudobulk glomeruli transcriptomes. Excel workbook with four tabs, one each for MN, FSGS and LN compared to CTRL and fourth tab that merges the three comparisons. Transcripts are labeled by official symbol and univariate T and F statistics are shown alongside effects sizes, p-values and BH adjusted p-values.

**Sup. Table 3.** – Differential expression analysis of pseudobulk glomeruli transcriptome and proteome. Excel workbook with two tabs, one for differential transcripts and one for differential proteins.

